# Cellular signatures of functional resilience in presymptomatic frontotemporal dementia

**DOI:** 10.1101/2025.02.24.25322778

**Authors:** Kamen A. Tsvetanov, Maura Malpetti, P. Simon Jones, Timothy Rittman, David J. Whiteside, Alexander G. Murley, Richard Bethlehem, Casey Paquola, Enrico Premi, Arabella Bouzigues, Lucy L. Russell, Phoebe H. Foster, Eve Ferry-Bolder, John C. van Swieten, Lize C. Jiskoot, Harro Seelaar, Raquel Sanchez-Valle, Robert Laforce, Caroline Graff, Daniela Galimberti, Rik Vandenberghe, Alexandre de Mendonça, Pietro Tiraboschi, Isabel Santana, Alexander Gerhard, Johannes Levin, Sandro Sorbi, Markus Otto, Maxime Bertoux, Thibaud Lebouvier, Simon Ducharme, Chris R. Butler, Isabelle Le Ber, Elizabeth Finger, Maria Carmela Tartaglia, Mario Masellis, Matthis Synofzik, Fermin Moreno, Barbara Borroni, Jonathan D. Rohrer, James B. Rowe, the Genetic FTD Initiative, GENFI

## Abstract

Frontotemporal dementia (FTD) shows autosomal dominant transmission in up to a third of families, enabling the study of presymptomatic and prodromal phases. Despite self-reported well-being and normal daily cognitive functioning, brain structural changes are evident a decade or more before the expected onset of disease. This divergence between cognitive function and brain structure contrasts with the coupling of structural and functional decline after symptom onset. In healthy ageing, it has been shown that functional connectivity is a better predictor of cognitive function than volumetric structural imaging. We previously proposed that in the presymptomatic phase of genetic FTD, the maintenance of brain functional network integrity enables mutation carriers to sustain cognitive performance. However, prior work has focused on a small number of, often predefined, networks. This provides a limited and potentially biased characterisation of the substrates and moderators of brain network integration. Here, we test the hypothesis that brain-wide functional integration in FTD determines resilience to progressive pathology before symptom onset. We assess functional connectome integration in 289 presymptomatic FTD-mutation carriers using functional magnetic resonance imaging in relation to cognition and contrast with 271 family members without mutations. Because structural atrophy, functional integration and cognitive profiles are multivariate, we used canonical correlation models, supplemented by multiple linear regression models for each imaging modality. We confirmed progressive atrophy and normal cognitive function in presymptomatic carriers compared to non-carriers. Notably, functional integration was preserved in presymptomatic carriers across age, while it declined in familial non-carriers. The strongest effects were observed in cognitive control networks. The changes in functional integration in presymptomatic carriers were behaviourally relevant and independent of the severity of atrophy, suggesting a resilience mechanism in those at risk of dementia. To generate hypotheses about the genetic and neurometabolic basis of resilience, we assessed the spatial overlap between behaviourally-relevant functional integration maps and gene transcription profiles. These spatial correlations suggested resilience signatures to glial cell composition (astrocytes, microglia, oligodendrocytes), revealing cellular mechanisms inaccessible to standard neuroimaging. Our findings suggest that resilience to atrophy arises from enhanced functional integration, protecting against clinical conversion for many years in individuals at risk of dementia. This result has implications for the design of presymptomatic disease-modifying therapy trials and gives hope for therapeutic strategies aimed at enhancing resilience and ability to maintain function despite the presence of genetically determined neuropathology.

## 1. Introduction

Neurodegeneration begins many years before the onset of symptoms in people at risk of dementia. Neuropathological and structural changes during this presymptomatic period are well established in Alzheimer’s disease, Huntington’s disease and frontotemporal dementia (FTD) using structural brain imaging, radiotracer imaging, and fluid biomarkers. However, it remains unclear why people remain functionally resilient to advancing neuropathology. We propose that the integration of functional network organisation during this presymptomatic stages confers resilience to the effects of neurodegeneration i.e. enabling cognitive function despite atrophy. The brain-wide spatial patterns of functional integration provides means to identify the mechanisms and the moderators of change in neural dynamics underlying cognition, including factors that mitigate the effect of neurodegeneration.

Maintaining cognitive abilities depends on the coupling of structural and functional properties of brain networks as people age (Geerligs et al., 2016; Liu et al., 2023; Persson et al., 2006). This phenomenon is amplified in those at risk of dementia (Rittman et al., 2019; Tsvetanov et al., 2020). The links between structure, function and performance have been influential in developing current models of neurocognitive ageing and dementia (Cabeza et al., 2018; Cope et al., 2018; Raj et al., 2012; Seeley et al., 2009). These associations contrast with the emerging evidence of neuropathological and structural changes many years before the onset of symptoms of Alzheimer’s disease, Huntington’s Disease and frontotemporal dementia (FTD) (Bonner-Jackson et al., 2013; Gregory et al., 2018; Kinnunen et al., 2018; Klöppel et al., 2015; Rohrer et al., 2015; Vatsavayai et al., 2016). This begs the question, why are some people resilient to pathology for so long?

Genetic FTD provides a model system to address this question. FTD has highly-penetrant gene mutations, providing the opportunity to define brain functional changes that facilitate maintenance of good cognitive function at the pre-symptomatic disease stage. Mutations in three main genes account for 10-20% of FTD cases: chromosome 9 open reading frame 72 (*C9orf72*), granulin (*GRN*) and microtubule-associated protein tau (*MAPT*). Phenotypic expression and age of symptom onset vary between (Deleon and Miller, 2018; Moore et al., 2020), but also within the same gene variant (Snowden et al., 2015), with environmental and secondary genetic moderation (Murphy et al., 2017; Premi et al., 2017). Nonetheless, all three mutations cause significant structural brain changes in key regions over a decade before the onset of symptoms in people who convert to symptomatic stages (Cash et al., 2018; Rohrer et al., 2015), as confirmed in longitudinal studies (Floeter et al., 2018; Jiskoot et al., 2019; Olm et al., 2018).

We recently proposed the maintenance of functional network integrity as one of the factors that enables presymptomatic mutation carriers to stay well in the face of progressive atrophy (Rittman et al., 2019; Tsvetanov et al., 2020). This connectivity-cognition relationship becomes stronger with age (Tsvetanov et al., 2016) and high risk of dementia (Liu et al., 2024b; Passamonti et al., 2019) – a finding generalised across modalities (e.g. MEG, Bruffaerts et al., 2019; Tibon et al., 2021; Tibon and Tsvetanov, 2022), cognitive states (Tomassini et al., 2022; Tsvetanov et al., 2018) and analytical approaches (Bethlehem et al., 2020; Geerligs et al., 2017; Liu et al., 2023). However, this work has focused on a small number of networks or predefined networks, which provides a limited and potentially biased characterisation of the substrates and the moderators of change in brain functional integration. This approach overlooks the distributed nature of cognitive substrates and neurodegenerative disease targets, which extend across multi-level, interactive networks, rather than being confined to single brain regions or networks (Margulies et al., 2016; Mesulam, 1998).

A common tool used to investigate functional networks and their connectivity in dementia is functional magnetic resonance imaging (fMRI). While functional connectivity from fMRI has faced challenges in understanding underlying mechanisms (Kullmann, 2020), these limitations can be addressed by linking functional measures to behavioural performance, modelling potential confounders (e.g. atrophy or systemic-to-vascular interactions), and employing data-driven approaches across the whole brain (Abdelkarim et al., 2019; Cabeza et al., 2018; Knights et al., 2024; Liu et al., 2023, 2024b; Tsvetanov et al., 2021b; Wu et al., 2022). Building on these advances, we propose a framework that identifies behaviourally-relevant, brain-wide patterns of functional integration and links them to gene transcription profiles (Arnatkevic̆iūtė et al., 2019; Hawrylycz and Jones, 2012). This approach not only overcomes traditional neuroimaging challenges but also reveals how these macroscopic neuroimaging phenotypes of functional resilience relate to normative spatial variations at cellular and molecular scales. This allows us to uncover insights into functional resilience that are typically inaccessible to standard neuroimaging techniques.

Here we sought to identify the brain-wide topology of network integration, providing means to identify the mechanisms and the moderators of change in neural dynamics underlying cognition to offset progressive neurodegeneration in the presymptomatic disease stage. This approach addresses the outstanding challenge that neither cognition nor the targets of neurodegenerative disease are mediated by single brain regions: they are distributed across multi-level and interactive networks. We therefore used a multivariate data-driven approach to identify differences in the multidimensional brain-behaviour relationship between presymptomatic carriers and non-carriers of mutations in FTD genes. We employed and extended a novel approach of multi-dimensional brain network analysis (Bethlehem et al., 2020) to define a brain-wide pattern of brain network organisation across levels of cortical organisation (Margulies et al., 2016; Mesulam, 1998), which are known to be differentially affected in frontotemporal dementia (Greaves and Rohrer, 2019; Migliaccio et al., 2024).

Our overarching hypothesis is that for those at genetic risk of dementia, the maintenance of higher-order cognitive networks confers cognitive reliance, preserving function despite pre-symptomatic structural degradation. We explore the spatial correspondence between brain-wide topology of network integration and normative regional variations in transcriptome-based cell-type distributions. We propose that the physiological and genetic signature of functional network resilience is shaped by the composition of cell-types essential for neuronal homeostasis under stress.

## 2. Materials and methods

### 2.1. Participants

The study design and the principal data processing pipelines are summarised in Figures 1 and 2. Twenty-five research sites across Europe and Canada recruited participants as part of an international multicentre partnership, the Genetic Frontotemporal Initiative (GENFI) (Cash et al., 2018; Rohrer et al., 2015). The study was given a favourable opinion by the Cambridge 2 Research Ethics Committee REC 17/EE/0032 IRAS ID 204052. Informed consent was obtained by all human participants. Inclusion criteria included anyone over the age of 18, who is symptomatic or a asymptomatic first-degree relative.

**Figure 1.**
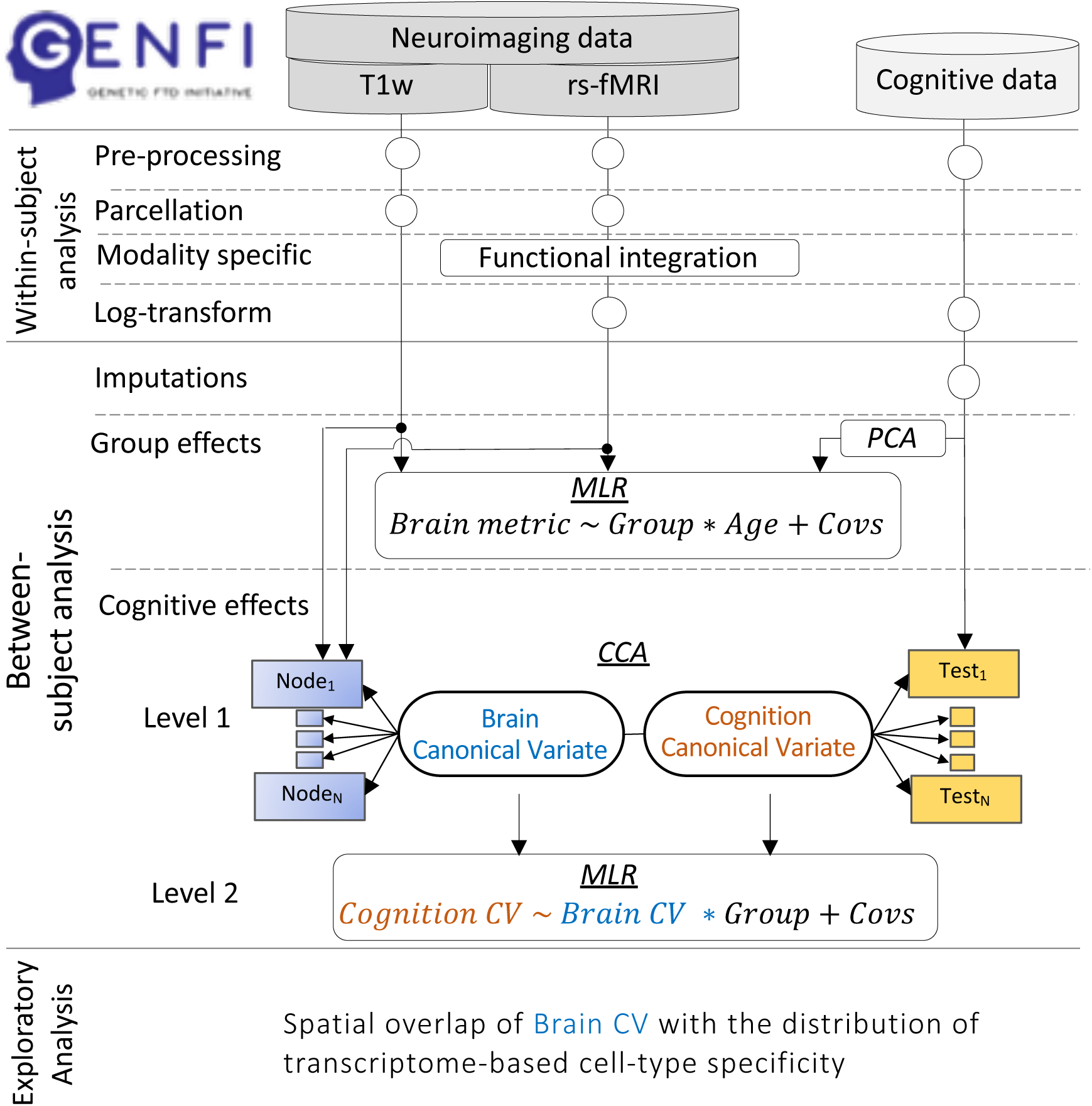
Schematic representation of modality datasets, their processing pipelines on a within-subject level, as well as data-reduction techniques and analytical strategy on between-subject level to test for group and cognitive effects of brain metrics. T1w, T1-weighted image acquisition; rs-fMRI, resting state functional magnetic resonance imaging acquisition; MLR – multiple linear regression; Group, individuals genetic status identity (i.e. presymptomatic carrier vs non-carrier); Covs, covariates of no interest; GLM, general linear model; CCA, canonical correlation analysis; CV, canonical variates from CCA analysis;

**Figure 2.**
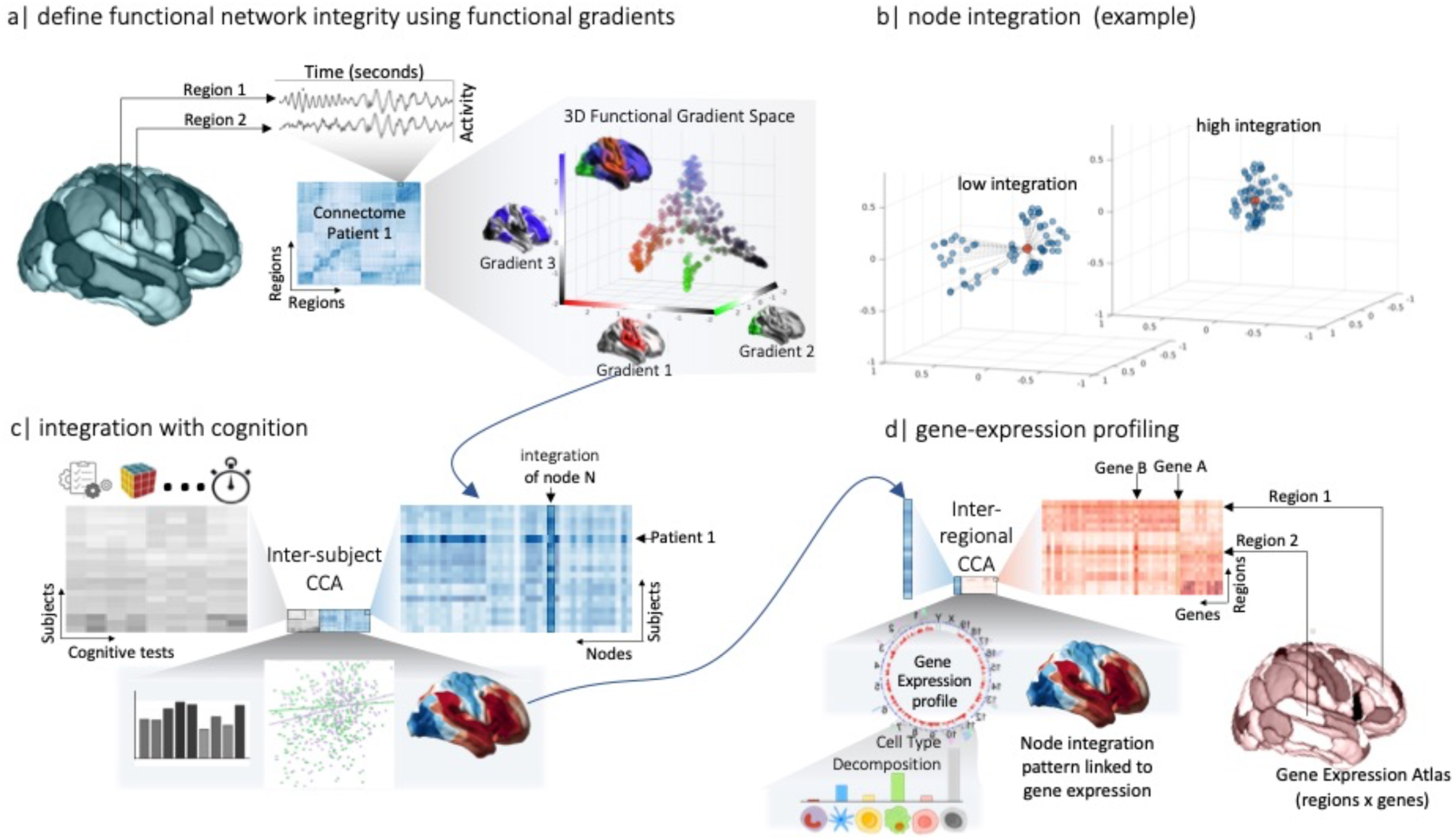
Schematic representation of defining network integration and its integration with cognition and gene-expression profiling. **(a) Functional network integration using functional gradient extraction:** Resting-state fMRI data is processed and parcellated into Glasser’s 360 nodes to estimate functional connectomes in terms of node-to-node functional connectivity. Individual estimates of the observed functional connectomes are concatenated as inputs to diffusion embedding approach. The first three gradients projected into a 3-dimensional gradient space and coloured by cortical surface colourmap above the scatter plot. Each circle indicates the location of a node in this 3D space with each functional gradient projected on the cortical surface next to the corresponding axis. Gradient 1 separates somatomotor and auditory cortex (red) from default mode regions (black). Gradient 2 extends between visual cortex (green) and default mode regions (black). Gradient 3 separates high-order regions (blue) from default mode and sensorimotor regions (black, red, green). The gradient space was constructed in a hold-out sample. **(b) Schematic illustration of the proposed integration metric** for a simulated node (red) and its community of nodes (blue) with low and high integration (left and right panels, respectively). Node integration is estimated as the inversed sum squared of Euclidean distances from each community node (blue) to the red node. Lower values represent a wider distribution of the community in the 3D gradient space, i.e. a more dispersed community (left panel). Higher values represent a denser distribution of the community, i.e. integrated community (right panel). The approach was repeated for each Glasser node in (a) for each individual’s own gradient space, where node community was based on the top 10% closest nodes in the hold-out template. **(c) Link between network integration and cognitive function:** individual estimates of functional integration and performance across a wide range of cognitive tests are concatenated (subjects by integration across regions and subjects by performance across cognitive tests) as inputs to inter-subject canonical correlation analysis (3, CCA). CCA identifies sources of signal in network integration and cognition that are related across individuals in terms of functional integration patterns and cognitive profiles and subjects scores. Second-level analysis tests whether the relationship between integration and cognition subject scores is stronger for presymptomatic carriers (evidence for functional resilience). **(d) Gene-expression profiling of functional integration**: the spatial correspondence between behaviour-relevant network integration map and gene expression data are integrated using inter-regional CCA. The gene expression profile expressed highly by functional integration map is analysed using Cell Type Decomposition, to identify cell-type enrichment based on the extent to which genes expressed the transcriptome map.

Participants were genotyped based on whether they carried a pathogenic mutation in *MAPT* or *GRN* genes or an expansion in c*9or*f72. Mutation carriers were classified as either symptomatic or presymptomatic based on clinical evaluation. Participants were classified as symptomatic if the clinician judged that symptoms were present, progressive and consistent with a diagnosis of a neurodegenerative disorder. A comparison group acted as controls, termed non-carriers, and comprised mutation-negative family members. In this study, based on DataFreeze 5, we focus on non-carriers (NC, N=271) and presymptomatic carriers (PSC, N=289) with usable structural and resting state functional magnetic resonance imaging data (MRI). Participants and site investigators were blinded to the research genotyping, although a minority of participants had undergone predictive genetic testing outwith the GENFI study. See Table 1 for demographic information and Table 2 for behavioural, cognitive and neuropsychological information of both groups.

**Table 1.**
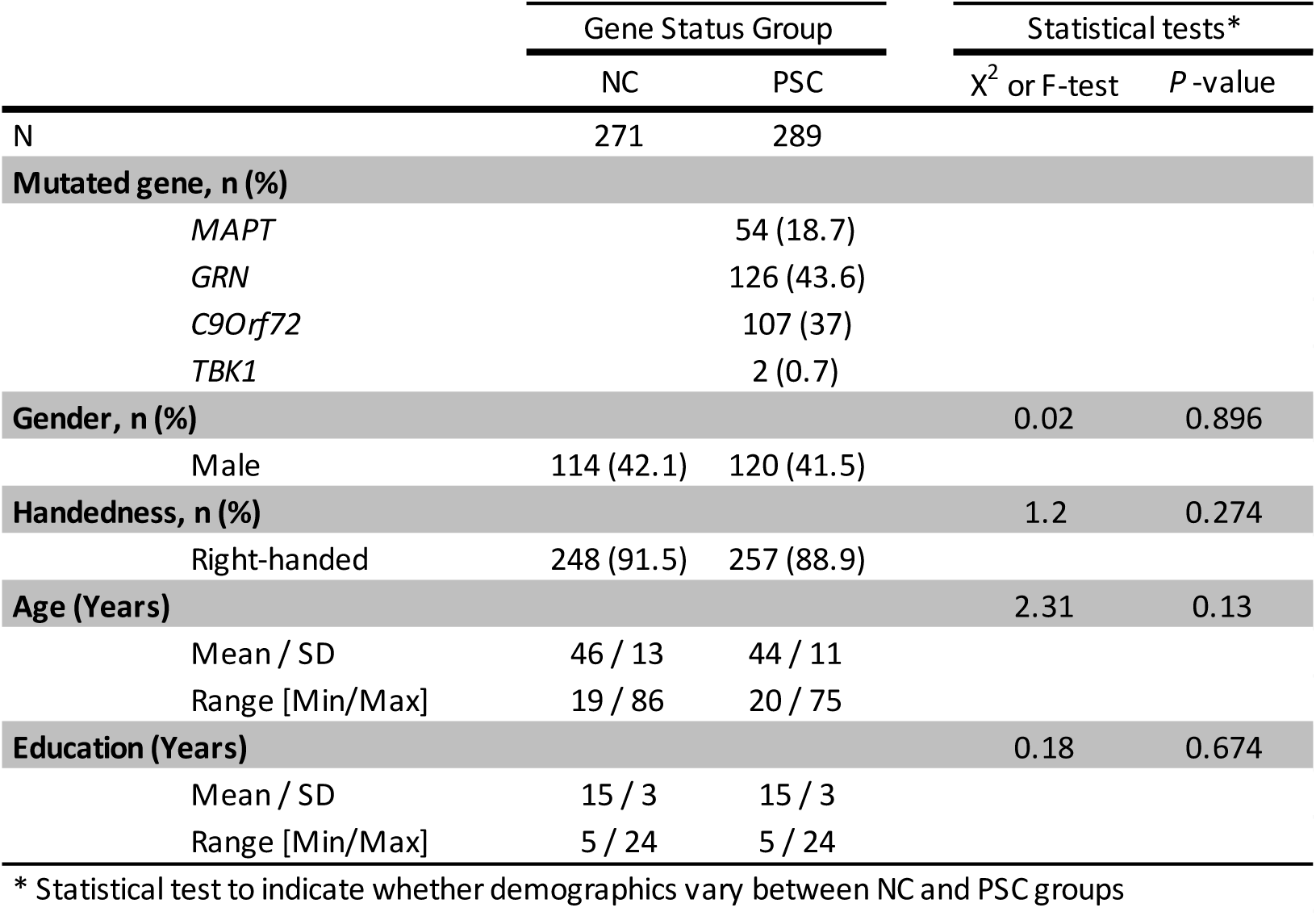
Demographics table with statistical test to indicate whether demographics vary between Non-Carriers (NC) and Pre-Symptomatic Carrier (PSC) groups.

|  | Gene Status Group |  | Statistical tests* |  |
| --- | --- | --- | --- | --- |
| | NC | PSC | $\chi^2$ or F-test | P-value |
| N | 271 | 289 |  |  |
| **Mutated gene, n (%)** |  |  |  |  |
| MAPT |  | 54 (18.7) |  |  |
| GRN |  | 126 (43.6) |  |  |
| C9orf72 |  | 107 (37) |  |  |
| TBK1 |  | 2 (0.7) |  |  |
| **Gender, n (%)** |  |  | 0.02 | 0.896 |
| Male | 114 (42.1) | 120 (41.5) |  |  |
| **Handedness, n (%)** |  |  | 1.2 | 0.274 |
| Right-handed | 248 (91.5) | 257 (88.9) |  |  |
| **Age (Years)** |  |  | 2.31 | 0.13 |
| Mean / SD | 46 / 13 | 44 / 11 |  |  |
| Range [Min/Max] | 19 / 86 | 20 / 75 |  |  |
| **Education (Years)** |  |  | 0.18 | 0.674 |
| Mean / SD | 15 / 3 | 15 / 3 |  |  |
| Range [Min/Max] | 5 / 24 | 5 / 24 |  |  |

\* Statistical test to indicate whether demographics vary between NC and PSC groups

**Table 2.** Average and standard deviation values with statistical test to indicate whether scores vary between Non-Carrier (NC) and Pre-Symptomatic Carrier (PSC) groups.

| Cognitive Test | Gene Status Group |  | Statistical tests* |  |
| --- | --- | --- | --- | --- |
| | NC | PSC | $\chi^2$ | P-value |
| Digit Span - Forwards | 7.7 ( 2) | 8 ( 2.1) | 3.14 | 0.076 |
| Digit Span - Backwards | 6.7 ( 2.2) | 6.9 ( 2.2) | 0.89 | 0.372 |
| Digit Symbol Task | 58 ( 14.3) | 57.5 ( 13.4) | 0.19 | 0.661 |
| Trail Making Test Part A | 27.7 ( 14.4) | 27.8 ( 10.6) | 1.71 | 0.192 |
| Trail Making Test Part B | 66 ( 33.2) | 67.8 ( 40.2) | 0.06 | 0.800 |
| Verbal Fluency - Letter | 41.2 ( 12.7) | 40 ( 14.3) | 1.23 | 0.268 |
| Verbal Fluency - Animal | 23.4 ( 5.8) | 24.1 ( 5.8) | 3.12 | 0.078 |
| Boston Naming Test | 31.8 ( 6.5) | 30.8 ( 6.8) | 0.15 | 0.697 |
| Block Design | 48 ( 15.1) | 47.6 ( 15.8) | 0.01 | 0.975 |
| FCSRT D Free | 12.2 ( 2.5) | 11.7 ( 2.9) | 2.44 | 0.118 |
| FCSRT Free | 31.8 ( 6.5) | 30.8 ( 6.8) | 1.75 | 0.186 |
| Stroop Color | 28.6 ( 5.8) | 29.7 ( 7.4) | 2.07 | 0.151 |
| Stroop Ink | 50.4 ( 12.6) | 52.6 ( 16.3) | 1.08 | 0.299 |
| Stroop Word | 22.5 ( 5.4) | 22.5 ( 5.6) | 0.01 | 0.983 |

### 2.2. Neurocognitive assessment

Each participant completed a standard clinical assessment consisting of medical history, family history, functional status and physical examination, in complement with collateral history from a family member or a close friend. In the current study 9 behavioural measures of cognitive function were correlated with neuroimaging measures. These included the Uniform Data Set (Morris et al., 2006): Digit Span forwards and backwards from the Wechsler Memory Scale-Revised, a Digit Symbol Task, Parts A and B of the Trail Making Test, the short version of the Boston Naming Test, and Category Fluency (animals). Additional tests included Letter Fluency and Wechsler Abbreviated Scale of Intelligence Block Design task, Free and Cued Selective Reminding Test (FCSRT) and Stroop tests. Latency measures for the Trail Making Test and Stroop tests were log-transformed and inverted so that higher values across all tests reflect better performance. Rate of missing neurocognitive data varied between 0 and 10%; hence, to increase statistical power and efficiency, missing data were imputed before further statistical analyses. Incomplete variables were imputed under fully conditional specification, using the default settings of the multivariate imputation by chained equations (MICE) in R (Buuren and Groothuis-Oudshoorn, 2011).

To test for differences in performance across cognitive measures, we performed principal component analysis as a widely used dimensionality reducing approach (Hotelling, 1933; Maaten et al., 2009; Pearson, 1901). For this purpose, we used Matlab’s function *pca.m* with default settings, where the number of components was determined using Horn’s parallel analysis (Dobriban, 2020; Horn, 1965) using 10,000 permutations. All variables were normalised to a mean of 0 and standard deviation of 1 prior to principal component analysis.

### 2.3. Neuroimaging assessment

#### 2.3.1. T1-weighted image acquisition and processing

Figures 1 and 2 provide a schematic representation of imaging data processing pipeline and the analysis strategy for linking brain-behaviour data. MRI data were acquired using 3T with optimised scanning protocols to maximise synchronisation across scanners and sites (Cash et al., 2018; Rohrer et al., 2015). A 3D-structural MRI was acquired on each participant using T1-weighted Magnetic Prepared Rapid Gradient Echo (MPRAGE) sequence over at least 283s (283-462s) and had a median isotropic resolution of 1.1mm, repetition time of 2000ms (6.6-2400), echo time of 2.9ms (2.8-4.6ms), inversion time of 8ms (8-9ms), and field of view 256×256×208mm. The imaging data were analysed using FSL pipelines (Jenkinson et al., 2012; Smith et al., 2004) and modules which called relevant functions from SPM12 (Friston et al., 2007). Native-space segmentation of GM, WM and CSF tissue and warps for normalisation to the Montreal Neurological Institute template space were estimated using FSL. In addition, anatomical images were processed and individually evaluated in FreeSurfer (Dale et al., 1999) Data were quality-control checked by semiautomated scripts monitored as previously described (Malpetti et al., 2020).

#### 2.3.2. fMRI image acquisition and processing

##### Preprocessing

For resting state fMRI measurements, Echo-Planar Imaging (EPI) data were acquired with at least six minutes of scanning. Analogous imaging sequences were developed by the GENFI Imaging Core team, and used at each GENFI study site to accommodate different scanner models and field strengths. EPI data were acquired over at least 308s (median 500s) and had a median repetition time of 2500ms (2200-2500ms), echo time of 30ms, flip angle of 80ms (80-85ms), in-plane resolution of 2.72×2.72mm (2.72-3.50 ×2.72-3.25), and slice thickness of 3.5mm (2.72-3.5).

To further ensure that potential group bias in head motion did not affect later analysis of connectivity, we took three further steps: i) fMRI data was further postprocessed using whole-brain Independent Component Analysis (ICA) of single subject time-series denoising, with noise components selected and removed automatically using *a priori* heuristics using the ICA-based algorithm (Pruim et al., 2015), ii) postprocessing of node-based time-series (see below) and iii) a subject-specific estimate of head movement for each participant (Jenkinson et al., 2002) included as a covariate in group-level analysis (Geerligs et al., 2017).

##### Post-processing and connectome estimation

For every individual, we projected ICA-denoised fMRI data to surface space and averaged vertex-based time-series within 360 nodes from a multimodal parcellation of the human cerebral cortex (Glasser et al., 2016). We post-processed nodal time-series to further reduce the effects of noise confounds on functional connectivity effects using the general linear model (Geerligs et al., 2017). This model included linear trends, expansions of realignment parameters, as well as average signal in WM and CSF, including their derivative and quadratic regressors (Satterthwaite et al., 2013) and CompCorr regressors (Behzadi et al., 2007) from the time-courses of each node. The average WM and CSF signals and CompCor signals were created by using the signal across all voxels with associated tissue type as defined by FSL’s segmentation. A band-pass filter (0.0078 to 0.1 Hz) was implemented by including a discrete cosine transform set in the GLM, ensuring the nuisance regression and filtering were performed simultaneously (Bright et al., 2017; Hallquist et al., 2013; Lindquist et al., 2019). Finally, the functional connectivity (FC) between each pair of nodes was computed using Pearson correlation on postprocessed time-series. Individual connectivity matrices were subjected to row-wise thresholding (top 10% of edges maintained) and converted into normalised angle matrices (Bethlehem et al., 2020; Margulies et al., 2016; Wael et al., 2020).

#### Gradient extraction

To determine the gradients of subject-level connectomes, we employed diffusion map embedding as a non-linear dimensionality reduction technique (Coifman and Lafon, 2006) shown to be highly reproducible (Hong et al., 2020), robust to confounds and sensitive to cognitive decline when compared to standard approaches of functional connectivity (Bethlehem et al., 2020).

First, a group-average functional connectivity matrix was constructed, thresholded, normalised and subjected to diffusion map embedding from an independent healthy adult cohort part of the Cambridge Centre for Ageing and Neuroscience, Cam-CAN, project (n=637) (Shafto et al., 2014; Taylor et al., 2015). The first three gradients explained 41.3% of the variance and were selected for further analysis. The gradients showed a functional differentiation running from sensory-to-transmodal abstract (G1), visual-to-transmodal abstract (G2) and somatomotor-to-transmodal executive (G3). This is in line with converging evidence that the transmodal cortex can be delineated into two sets of regions (Margulies and Smallwood, 2017; Vatansever et al., 2017; Vidaurre et al., 2017): (i) regions hypothesized to serve more abstract functions, e.g. default mode network (Raichle, 2015; Raichle et al., 2001) and (ii) regions hypothesized to serve intelligence and executive control (Assem et al., 2020; Duncan, 2010; Woolgar et al., 2018; Wu et al., 2022). Together, these gradients describe functional discrimination between sensory modalities and across levels of the cortical hierarchy (i.e. sensory processing, abstract thinking and higher-order cognition) which are known to be differentially affected in frontotemporal dementia (Greaves and Rohrer, 2019; Migliaccio et al., 2024). Regional gradient values reflect the expression of connectivity profiles in relation to that axis (e.g.: two regions with similar gradient values exhibit a similar distribution of rs-FC along the sensory-abstract transmodal axis).

The gradient template generated in the CamCAN cohort was used as a reference to which each GENFI individual gradient maps were aligned using Procrustes transformation (Wang and Mahadevan, 2008). The degree of deformation and translation of transformation matrix for an individual’s gradients to align to the gradient template was used to evaluate potential bias in the alignment procedure.

##### Node integration using multi-dimensional gradients

To investigate differences in multi-dimensional cortical organisation, we quantified a new metric, termed here *‘node integration’*. This metric aimed to provide a more granular representation of previously proposed metric of network integration in this multi-dimensional functional connectivity space (Bethlehem et al., 2020) across seven large-scale networks (Yeo et al., 2011). This allowed us to estimate a measure of integration at the regional level, rather than integration measures at the network-level. Here, we focus on the first three gradients, forming a 3D space where each axis reflects the values along each gradient. We calculated ‘node integration’ as the inverse of the sum squared of Euclidean distances from each community’s centre of gravity to each node in that community (i.e. moving from large-scale networks in Bethlehem et al, to communities, allowing us to focus on node integration within segregated subsystems of the brain). The node community was based on the top 10% closest nodes in the group average such that high values indicate high integration (Figure 2b, top scatter plot) and low values indicate low integration i.e. high dispersion (Figure 2b, bottom scatter plot). The approach was repeated for each node in each participant’s own gradient space. For completeness, we also explore the consistency of effects across varying the thresholds (e.g. 5% vs 10% vs 15% vs 20%).

### 2.4. Statistical analysis

Statistical analysis used Matlab 2020b calling the packages as described below. We performed a descriptive analysis of all the characteristics from demographic, clinical and cognitive characteristics within the sample before integrating these with neuroimaging data as described below.

#### 2.4.1. Group and progression effects in brain structure and function

To determine gene-carrier group effects and pre-disease progression effects on each brain metric (cortical thickness and functional integration) of each node we used a robust linear regression as implemented in Matlab’s function “fitlm.m”. Gene-carrier group effects were modelled based on group identity (presymptomatic carrier vs non-carrier) based on whether an individual was a carrier of a pathogenic variant in c9orf72, MAPT or GRN. Pre-disease progression effects were modelled using the interaction between group identity and individual’s age, on the basis that the pre-disease progression process in presymptomatic carriers differs from the normal ageing process in non-carriers. GENFI’s large-sampled cohort was created using harmonized multi-site neuroimaging data. Although, scanning protocols were optimised to maximise comparability across scanners and sites (Cash et al., 2018; Rohrer et al., 2015), different scanning platforms can introduce systematic differences which might confound true effects of interest (Chen et al., 2014). Therefore, in the analysis of neuroimaging data we included scanner site in addition to sex and head motion (Power et al., 2012) as additional covariates of no interest. The regression model was specified by Wilkinson’s notation (Wilkinson and Rogers, 1973), *‘Brain metric ∼ group*age + sex + head motion + scanner site’* (where group is the gene status: mutation carrier vs non-carrier) and fitted for each node separately. We tested the same model with *cognitive function* as a dependent variable. To test whether the effects of functional integration are independent of atrophy effects additional model was performed including cortical thickness of the corresponding node as a covariate of no interest (i.e. *‘Functional integration ∼ group*age + sex + head motion + scanner site + cortical thickness’*). Models were evaluated using 10,000 permutation tests and reported at p-perm<.05. A sensitivity analysis was performed using a multivariate approach, which is described in the next section.

#### 2.4.2. Brain-behaviour relationships

For brain-behaviour analysis, we adopted a two-level analytical procedure (Passamonti et al., 2019; Tsvetanov et al., 2022, 2020, 2018, 2016). In the first-level analysis, we assessed the multidimensional brain-behaviour relationships using regularized canonical correlation analysis with permutation-based 10-fold cross-validation. This analysis described the linear relationships between the two multivariate sets of variables, namely node integration and behavioural performance, by providing pairs of canonical variates (Node Integration-CVs and Cognition-CVs) as linear combinations of the original variables which are optimised to maximise their correlation. Namely, dataset 1 consisted of node integration across 360 nodes (i.e. Node integration dataset). Dataset 2 included the performance measures on the nine tests (i.e. Cognition dataset). Next, we tested whether the identified behaviourally-relevant CVs of node-integration were differentially expressed by NC and PSC as a function of age. To this end, we performed a second-level analysis using robust linear regression. Independent variables included subjects’ node integration scores from first level CCA, group identity and their interaction term (e.g. brain scores x group). The dependent variable was subjects’ cognitive scores from the first level CCA (Cognition-CV). Given that the interaction effects were derived from continuous variables, we tested and interpreted interactions based on simple slope analysis and slope difference tests (Aiken and West, 1991; Dawson, 2014; Dawson and Richter, 2006). Covariates of no interest included age, sex, head motion and site scanner.

#### 2.4.3. Spatial covariance of behaviour-relevant node integration with tran-scriptome-based cell-type decompositions

To generate hypotheses about the genetic and neurometabolic basis of resilience, we assessed the spatial overlap between behaviourally-relevant functional integration map (Function-CV regional loadings) and cell-type parameters using gene transcription profiling maps. Spatial covariance between node integration map and gene expression across 9394 genes expressed in the human brain (Hawrylycz and Jones, 2012) was based on CCA association using 10,000 spin-permutation-based 10-fold cross-validations (Tsvetanov et al., 2022, 2020). The spin-based permutations (p-spin) aimed to preserve spatial autocorrelation (Alexander-Bloch et al., 2018; Fulcher et al., 2021; Váša and Mišić, 2022). The latent variable represented a spatial pattern of gene expression that covaried significantly with the node integration pattern associated with cognitive function. Full details about the processed transcriptomic data are available elsewhere (Arnatkevic̆iūtė et al., 2019). Genes highly expressing this pattern (i.e. high loadings) were tested against a molecular atlas of human brain vasculature of 17 control and Alzheimer’s disease patients (Yang et al., 2022) using cell-type decomposition (Hansen et al., 2021). This enabled us to test whether the resilience-relevant genes were preferentially expressed in specific cell types, i.e., testing for gene sets specific to eleven major canonical cortical cell classes: astrocytes (Astro); brain endothelial cells (BEC); ependymal cells (Epend); macrophage/microglia (MacMic); meningeal fibroblasts (MFibro); neurons (Neuro); oligodendrocyte precursors (Opc); oligodendrocytes (Oligo); pericytes (Peri); perivascular fibroblasts (PFibro); and smooth muscle cells (SMC). To this end, we calculated the ratio of genes in each set preferentially expressed by each cell type (e.g. ratio for pericytes is calculated from the number of genes preferentially expressed in pericytes divided by the total number of genes). Gene sets were thresholded to include the top *n*% of genes with significant loadings, where significance threshold varied from 0.001 to 1 (all genes). Statistical significance was determined using a null distribution of ratios based on 10,000 sets of random genes (Hansen et al., 2021).

### 2.5. Data and code availability

Data were acquired from GENFI data freeze 5. Anonymized data not published within this article will be made available by request from any qualified investigator and can be requested via the GENFI website (https://www.genfi.org/contact-us-2) or via Dementias Platform UK (https://portal.dementiasplatform.uk/Apply).

Code and composite data to reproduce manuscript figures and statistical analyses are available at https://github.com/kamentsvetanov/functional_resilience_ftd. Surface-based analysis of anatomical images were performed in FreeSurfer (Dale et al., 1999). Resting-state fMRI data were pre-processed using FSL pipelines (Jenkinson et al., 2012; Smith et al., 2004) and modules which called relevant functions from SPM12 (Friston et al., 2007). Post-processing was based on a GLM-like approach (Geerligs et al., 2017) available at https://github.com/MRC-CBU/riksneurotools/blob/master/GLM/. Visualisation of neuro-imaging results was in MRIcroGL and BrainSpace, while ridgeline plots generated using R-based ggridges and ggplot2 (<u>10.32614/CRAN.package.ggridges</u>) (R Core Team, 2021; Wickham, 2016). Spin permutations used code available at https://github.com/frantisekvasa/rotate_parcellation. Fully-pre-processed transcriptomic data were available at https://figshare.com/articles/dataset/AHBAdata/6852911 and https://github.com/BMHLab/AHBAprocessing. The molecular atlas of the human brain vasculature was made available previously (Yang et al., 2022). Code for cell-type decomposition analysis is available at https://github.com/netneurolab/hansen_genescognition.

## 3. Results

### 3.1. Participants

Characteristics of the presymptomatic carriers and non-carriers in the current GENFI sample are detailed in Table 1. In a group of 289 presymptomatic carriers, 18.7% were *MAPT* mutation carriers, 43.6% were *GRN* mutation carriers, 37% were *C9Orf72* mutation carriers and 0.7% were *TBK1* mutation carriers. Among the presymptomatic carriers, nearly 89% of presymptomatic carriers were right-handed, 42% were male, the average age was 44 years and the average number of years spent in education was 15 years. Presymptomatic carriers did not differ from non-carriers across these demographics variables, Table 1, and individual cognitive tests, see Table 2. The percentage of missing values across cognitive tests varied between 0 and 10%. In total of 560 participants, 12 records were incomplete for Trail Making datasets; 2, 3 and 14 records were incomplete for Digit Symbol, Boston Naming and Block Design datasets, respectively; 4 and 6 records were incomplete for Fluency Animals and Fluency Letter data; and 56 and 49 records were incomplete for FCSRT and Stroop data, respectively.

### 3.2. Group differences

#### Cognitive function

Cognitive function was represented by a single principal component, *P*<.001, explaining 39.5% of the total variance across cognitive measures and reflecting overall cognitive function (Figure 3). The multiple linear regression model testing for overall group differences and group-age interaction in the cognitive function component between presymptomatic and non-carriers were not significant (r=-.07, *P*=.084 and r = -.05, *P*=.175). This indicates that cognitive function levels in presymptomatic carries are comparable to non-carriers, as expected.

**Figure 3.**
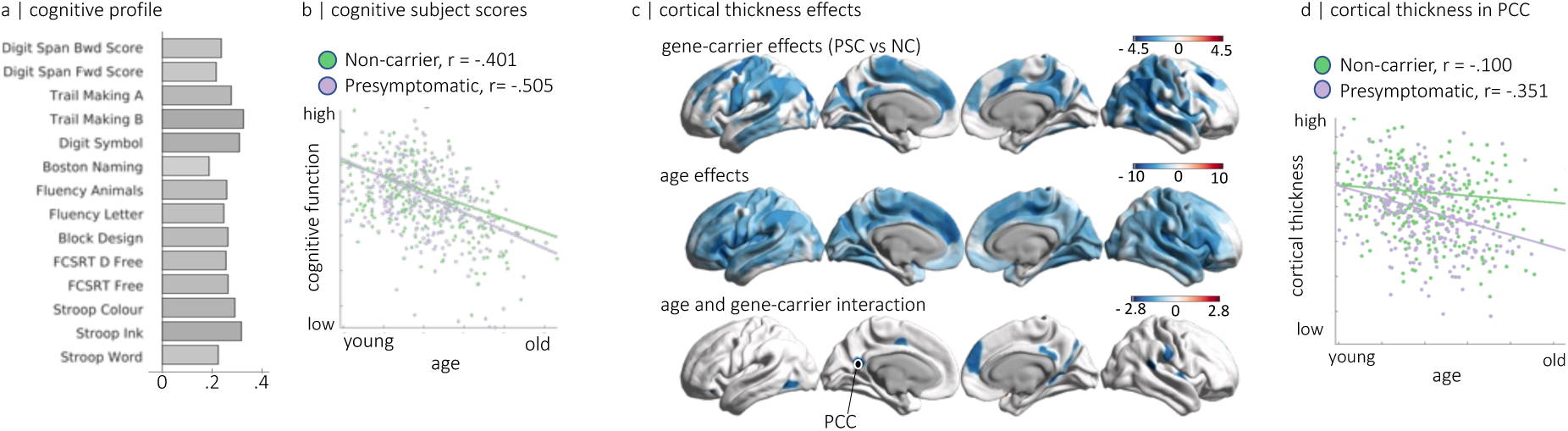
Cognitive function and cortical thickness effects. a| Cognitive profile for first, and only, significant principle component across cognitive measures. (b) scatter plot of cognitive subject scores, visualising the relationship between age and cognitive performance in each genetic status group separately. Each point indicates the strength to which an individual expresses the cognitive profile. (c) Top panel, spatial visualisation of cortical thickness effects for group differences between presymptomatic carriers (PSC) and non-carriers (NC), where the cold colour scheme indicates the strength of effect size of pre-symptomatic carriers (PSC) showing less cortical thickness than non-carriers (NC), while the hot colour scheme indicated the opposite effect (i.e. PSC > NC). Middle panel, age-related effects of cortical thickness. Bottom panel, age and group interaction effects with cold colour scheme indicating disproportionately stronger age effect in PSC than in NC (i.e. disease progression). (d) scatter plot of region-specific effects (PCC as indicated by a black circle in bottom panel of (c)) visualising the relationship between age and cortical thickness in each genetic status group, separately. The ticks on the x-axis in scatter plots were set from -2 to 3, in increments of 1, while the ticks on y-axis ranged from -3 to 2, also in steps of 1.

#### Cortical thickness

The multiple linear regression model testing for overall group differences in cortical thickness between presymptomatic and non-carriers was significant in frontal, temporal, parietal and subcortical regions (Figure 3), reflecting expected presymptomatic differences in brain-wide atrophy. Beside a strong brain-wide age-related differences in cortical thickness, a small set of regions, including anterior and posterior cingulate and superior temporal gyrus, showed a significant interaction effect of age and gene status (Age:Gene Status) (Figure 3c). The interaction effect reflected a stronger age-related decrease in cortical thickness for presymptomatic carriers versus non-carriers (i.e. disease progression of atrophy effects in presymptomatic FTD), Figure 3d. Overall, the observed univariate effects were consistent with the multivariate analysis using canonical correlation analysis (Figure S3).

#### Functional integration

The multiple linear regression test for group differences in functional integration was significant in several areas across the frontal cortex, including inferior frontal gyrus and anterior cingulate (Figure 4a). These effects were in the presence of strong age-related decrease in functional integration (Figure 4b). There was a significant interaction between age and gene status (Age:Gene Status) in regions classically associated with cognitive control (anterior cingulate, dorsolateral prefrontal cortex and intraparietal sulcus), and the default mode (ventromedial cingulate, posterior cingulate and lateral temporal pole). The interaction effects were driven by a strong decrease in functional integration in non-carriers with age *versus* minimal age effects in presymptomatic carriers i.e. presymptomatic carriers sustain functional integration more than non-carrier peers. Sensitivity analyses confirmed that these effects were robust to varying levels of functional community thresholding (Figure S1), atrophy levels (Figure S2), and multivariate analysis (Figure S3).

**Figure 4.**
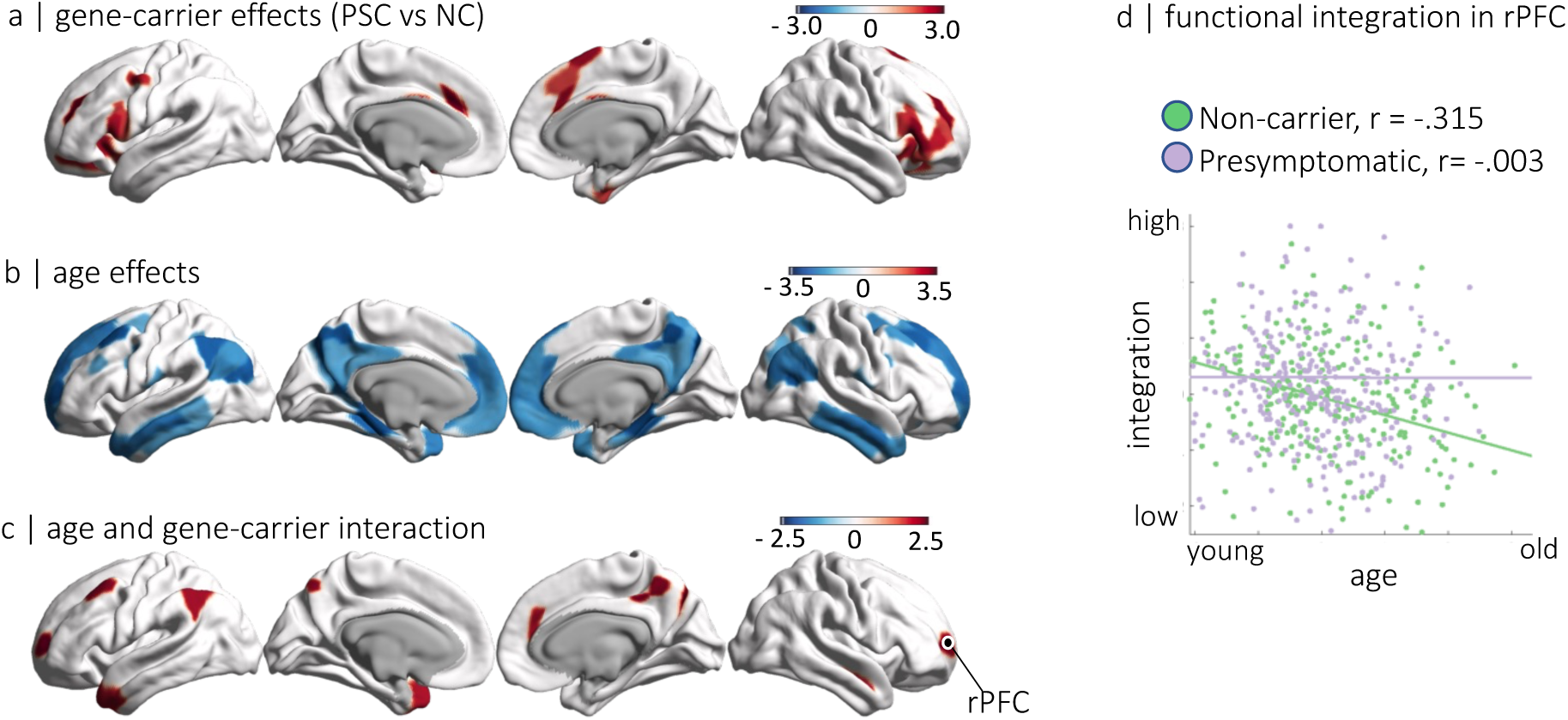
Functional integration effects. (a) Group differences between presymptomatic carriers (PSC) and non-carriers (NC), where the hot colour scheme indicates the strength of effect size of pre-symptomatic carriers (PSC) showing higher functional integration than non-carriers (NC), while the cold colour scheme indicated the opposite effect (i.e. PSC < NC). (b) Age-related effects of functional integration with cold colours showing age-related decrease in functional integration. (c) age and group interaction effects with hot colour scheme indicating disproportionately stronger age effect in NC versus PSC (i.e. functional resilience in PSC). Maps are thresholded at p-perm<.05, based on 10.000 permutations. For more complete description of the spatial representations see un-thresholded maps in Figure S1b. (d) scatter plot of region-specific effects (rPFC as indicated by a black circle in c) visualising the relationship between age and functional integration in each genetic status group, separately. The ticks on the x-axis were set from -2 to 3, in increments of 1, while the ticks on y-axis ranged from -3 to 2, also in steps of 1.

### 3.3. Brain-behaviour relationships

#### Cortical thickness and cognition

Canonical correlation analysis of cortical thickness and cognition identified one significant pair of canonical variates (r=.32, *P*<.001). Variable loadings and subject scores reflecting the strong positive relationship between cortical thickness and cognitive performance are shown in Figure 5. The cortical thickness canonical variate (Thickness-CV) expressed high loadings in frontal (dorsal-lateral prefrontal gyrus, precentral gyrus, paracentral lobule), parietal (postcentral gyrus, superior and inferior parietal lobule), superior temporal gyrus and occipital (lateral and medial occipital cortex) regions (Figure 5). The Cognition-CV profile expressed positively a large array of cognitive tests, with strongest values on Block design, Trail Making, Digit Symbol, Selective Reminding tests and Digit Span. The positive correlation between cortical surface and cognitive CV’s confirms the expected brain-behaviour relationship across the cohort as a whole, between cortical thickness and attention, processing speed, visuospatial ability and working memory.

**Figure 5.**
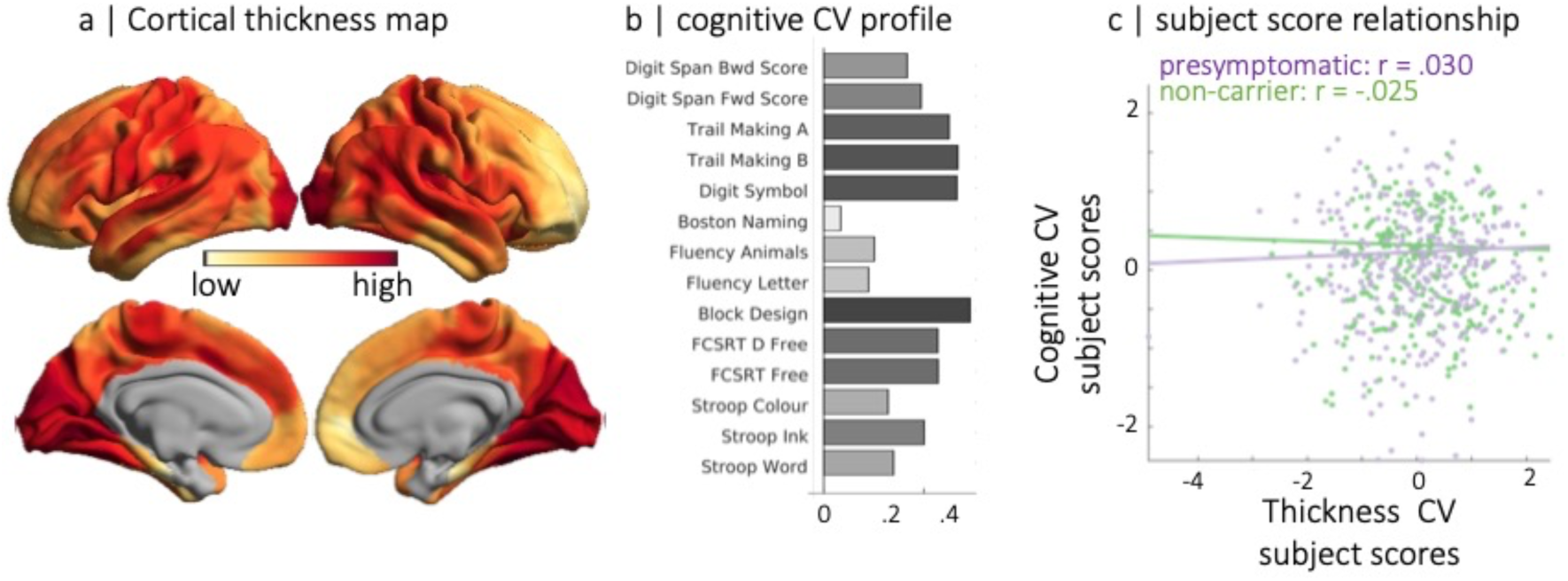
Link between cortical thickness and cognition using canonical correlation analysis. (**a**) Spatial distribution of parcellated cortical thickness value (i.e. loading values ranging from -.085 to 0.42) where light yellow to dark red colours are used for the strength of positive correlation with cognitive function profile in panel **b**. (**c**) The scatter plot represents the relationship (or lack thereof) between cortical thickness and cognition subjects scores for presymptomatic carriers (purple) and non-carriers (green) after accounting for age and other covariates.

To understand the structure-cognition relationship in each group and in relation as a function of age, we performed a second-level interaction analysis using a regression model: We entered Cognition-CV subject scores as dependent variable, and Thickness-CV subject scores, genetic status (i.e., mutation carrier or non-carrier), age, and their interactions as independent variables in addition to covariates of no interest (see Materials and methods). The results indicated that the relationship between cortical thickness and cognition no longer holds after including age and other covariates in the model (Figure 5). This suggests that the link between cortical thickness and cognition identified by CCA merely reflects age and/or other covariates of no interest in the model. This aligns with research indicating that structural brain-behaviour correlations are often difficult to establish (Boekel et al., 2015; Raz and Rodrigue, 2006), suggesting that brain structure alone is insufficient to account for the complex relationship between neural processes and observable behaviours.

#### Functional integration and cognition

CCA of functional integration and cognition identified two significant pairs of canonical variates (Function-CV1 and Cognition-CV1, r=.15, P=.036; Function-CV2 and Cognition-CV2, r=.19, P=.005). Variable loadings and subject scores reflecting the strong positive relationship between functional integration and cognitive performance are shown in Figure 6. The Function-CV1 expressed high functional integration values in frontal (lateral prefrontal cortex and anterior cingulate), intraparietal regions and inferior temporal regions. The Cognition-CV1 expressed positively Digit symbol, Block Design, Digit Span and Trail making tests. This first canonical variate pair indicated that higher performance on higher cognitive function tests, involving working memory, visuospatial reasoning and processing, and problem-solving is associated with stronger functional integration in the frontoparietal network.

**Figure 6.**
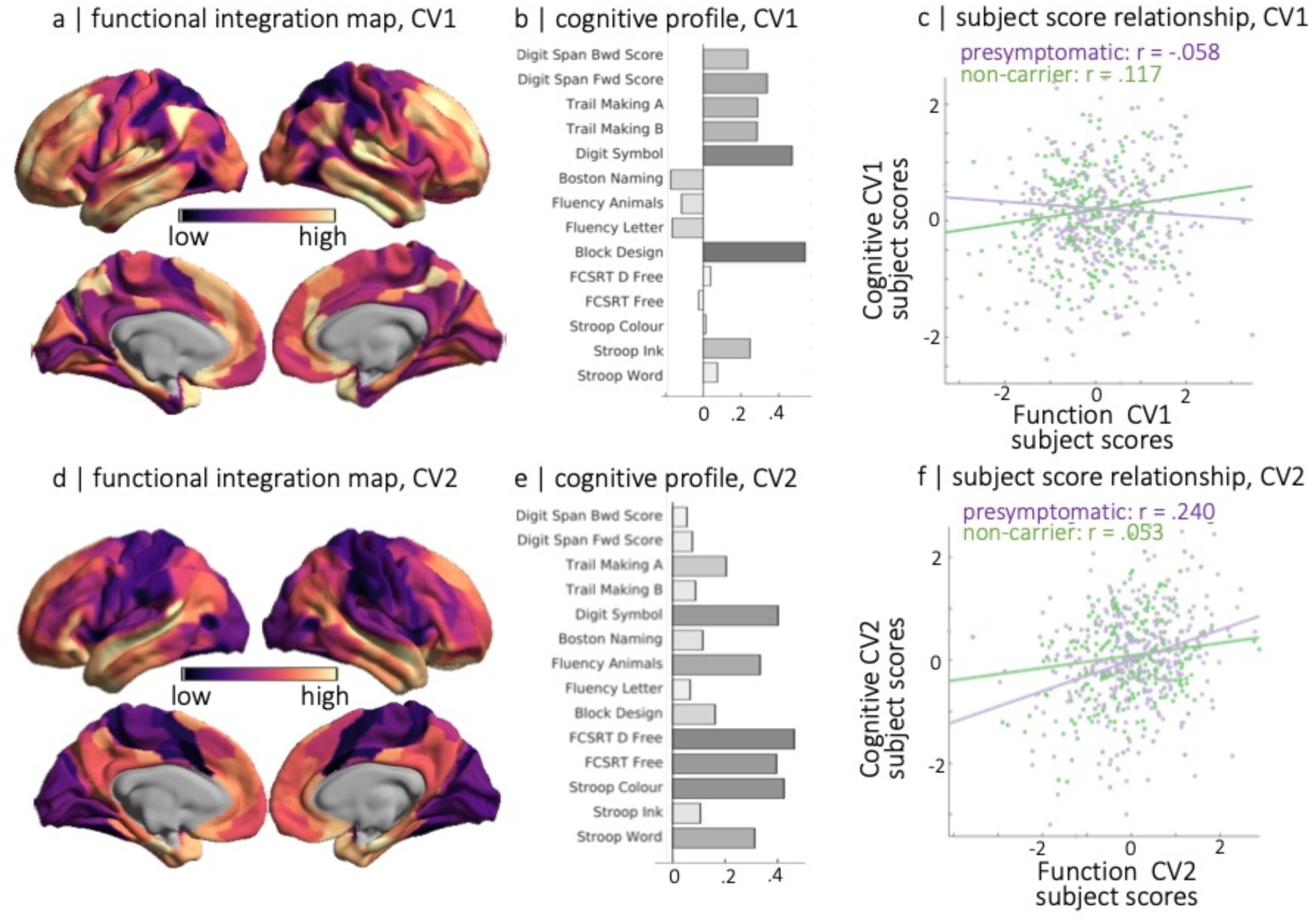
Link between functional integration and cognition using canonical correlation analysis. (**a**) Spatial distribution of functional integration nodal values for Canonical Variate 1 where dark purple to light yellow colours are used for the strength of positive correlation with cognitive function profile in panel **b**. (**c**) The scatter plot represents the positive relationship between functional integration and cognition subjects scores for presymptomatic carriers (purple) and non-carriers (green) after accounting for age and other covariates. (d-f) represent results as shown in (a-c) for Canonical Variate 2.

The Function-CV2 expressed high functional integration values in frontal (medial frontal cortex, inferior frontal gyrus), parietal (precuneus, intraparietal gyrus), superior temporal regions. The Cognition-CV2 expressed positively all tests, with strongest loading values on Digit Symbol, Fluency, Selective Reminding and Stroop tests. This second canonical variate pair indicated that higher performance on a wide range of cognitive tests, involving memory, attention and mental processing speed is associated with stronger functional integration.

To test whether the observed behaviourally-relevant pattern of functional integration is differentially expressed between genetic status groups and age, we constructed a second-level regression model with robust error estimates by including Function-CV subject scores, genetic status, age, and their interaction terms as independent variables and Cognition-CV as dependent variable in addition to covariates of no interest.

We found evidence for significant interaction between genetic status and Function-CV1 (r=-.09, *P*=.019) explaining unique variance in Cognition-CV1. We used simple slope analysis and slope difference tests (Aiken and West, 1991; Dawson, 2014; Dawson and Richter, 2006) to test formally for differences in the relationship between Function-CV1 and Cognition-CV1 for presymptomatic carriers and non-carriers. The relationship between Function-CV1 and Cognition-CV1 was stronger for non-carriers relative to presymptomatic carriers (r=.09, *P*=.019), indicating the increasing importance of functional integration in frontoparietal network to maintain higher cognitive order function across the health lifespan, Figure 6c.

For Canonical Variate 2, the interaction between genetic status and Function-CV2 (r=.10, *P*=.019) and Function-CV2 (r = .15, *P* < .001) explained unique variance in Cognition-CV2. Simple slope analysis and slope difference tests revealed that the relationship between Function-CV2 and Cognition-CV2 was stronger for presymptomatic carriers relative to non-carriers (r=.09, *P*=.025), indicating the increasing importance of functional integration in medial frontal and temporoparietal regions for presymptomatic carriers to maintain performance, Figure 6f.

### 3.4. Spatial overlap of brain functional resilience maps with brain cell-type distributions

We next used normative transcriptomics to identify genes that are normally preferentially expressed in the regions associated with brain functional resilience in presymptomatic frontotemporal dementia (i.e. Function-CV2). Regularised-CCA identified one latent component (r=0.58, p=0.001, Figure 7). Using cell-type decomposition analysis on vascular cell-type specific gene sets, we determined the ratio of genes in each gene set preferentially expressed across eleven cortical cell types. Gene-sets were thresholded to include the genes with significant loadings at p-value<.05 (10,000 permutations) (Figure 7). The results were consistent across various thresholds and labelling schemes for cell-type decomposition (Figure S4). Highly ranking genes were significantly more expressed in glial cells (astrocytes, macrophage/microglia and oligodendrocytes) and significantly less expressed in brain endothelial cells. Broadly, we find evidence that areas associated with functional resilience in presymptomatic frontotemporal dementia are enriched for expression of genes related to glial cell function and neuroinflammation, and underexpressed for genes in endothelial cells.

**Figure 7.**
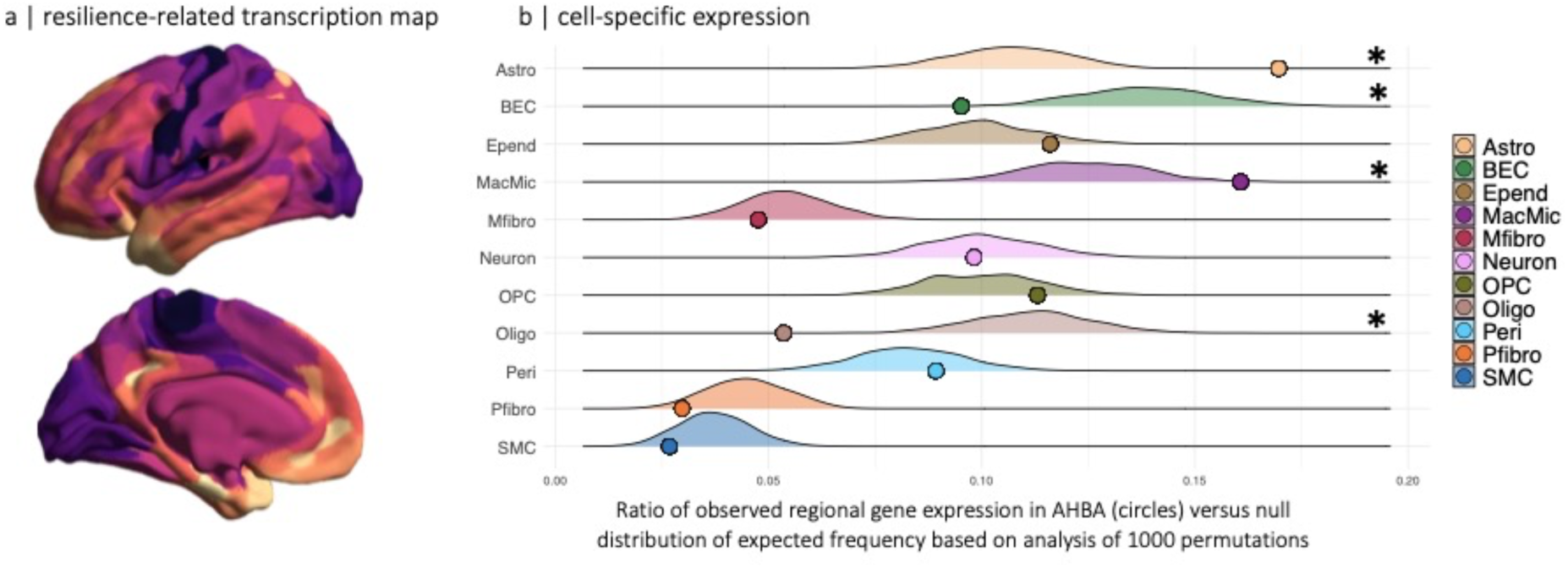
Spatial correspondence between functional resilience map expressed by +Canonical Variate 2 and cell-type decomposition. (a) Spatial map of the weighted whole genome expression profile correlated with the resilience-induced functional integration map. (b) Cell-type decomposition was used to identify cell-type enrichment based on extent to which genes expressed the transcriptome map in a. Gene sets for each cell-type were constructed by thresholding of genes with significant loadings, p-value < 0.05. Note that results were consistent across a range of thresholds and labelling schemes for cell-type decomposition, see Figure S5. The ratio of genes in each gene set preferentially expressed in eleven distinct cell-types (circles) is shown against their null distribution of a model with random selection of all genes (10,000 permutations, *p-value < 0.05). For example, pericyte’s ratio is calculated from the number of genes preferentially expressed in pericytes divided by the total number of genes. Cell type-specificity of genes is described elsewhere (Yang et al., 2021) Astro – astrocytes, BEC – brain endothelial cells, Epend – ependymal, MacMic – macrophage/microglia, Mfibro – meningeal fibroblast, Neuron – neuron, OPC – oligodendrocyte precursor cells, Oligo – oligodendrocytes, Peri – pericytes, Pfibro – perivascular fibroblast, SMC – smooth muscle cells.

## 4. Discussion

We have identified brain-wide differences in structural and functional imaging between familial non-carriers and presymptomatic carriers of frontotemporal dementia-related genetic mutations. Despite observing atrophy patterns reflecting disease progression, cognitive performance remained similar between groups. Functional node integration in cognitive control networks was preserved in carriers across age, suggesting a resilience mechanism in those at risk of FTD. This resilience was further evidenced in brain-cognition relationships: while grey matter-cognition relationships were consistent across groups, presymptomatic carriers showed stronger associations between functional integration and cognition. The spatial profile of functional resilience corresponded with normative variations in cell-type distributions, indicating that enhanced glial support may underlie this resilience. These findings offer new insights into the resilience mechanisms underpinning the complex interplay between structural atrophy, functional adaptations, and cognitive preservation in presymptomatic frontotemporal dementia.

People with genetically determined high risk of dementia can maintain good cognitive abilities and successful daily functioning despite significant neuronal loss during presymptomatic stages. The ability to maintain functional integration in the presence of neuropathology suggests a form of resilience (Cabeza et al., 2018; Stern et al., 2020), which may play a part in delaying symptoms onset and preserving quality of life in individuals at risk of FTD. The dissociation between structural decline and preserved function can occur to some extent with healthy ageing (Liu et al., 2023, 2022) but is exaggerated in presymptomatic genetic FTD as shown in the current study.

While earlier work on resilience in FTD and related disorders typically focused on global measures or a limited set of brain regions (Liu et al., 2024a; Rittman et al., 2019; Tsvetanov et al., 2020), we identified the brain-wide spatial pattern of functional resilience. This comprehensive coverage is an advantage, as neither the functional anatomy of cognition nor the foci of neurodegeneration are confined to singular regions, but rather are distributed across multi-level, interactive networks (Margulies et al., 2016; Mesulam, 1998). To identify the whole brain topology of resilience, we developed a novel multi-dimensional brain network approach (Bethlehem et al., 2020). This delineates the brain-wide pattern of network organization across levels of cortical hierarchy and in relation to cognitive preservation. This methodology offers three key advantages.

First, the brain-wide patterns of functional resilience can be used to identify potential mediators and moderators of resilience by linking receptor/metabolic imaging templates (Hansen et al., 2022; Markello et al., 2022) and gene transcription profiles (Arnatkevic̆iūtė et al., 2019; Hawrylycz and Jones, 2012). Our cell-type specific gene expression analysis highlighted the resilience-relevance of genes preferentially expressed in glial cells (astrocytes, macrophage/microglia and oligodendrocytes) and under expressed in endothelial cells (Hansen et al., 2021; Yang et al., 2022). Glial cells are involved in neuronal homeostasis and defence mechanisms against disease conditions, including physiological neuroplasticity, synapse turnover, brain barriers health and protection against oxidative stress (Verkhratsky and Butt, 2023; Verkhratsky and Pivoriūnas, 2023). This is well aligned with the idea that glial cells are intricately related to cognitive resilience (Verkhratsky and Zorec, 2024). Our findings motivate further understanding of the molecular processes underlying defence responses to neurodegeneration and for targeted interventions supporting resilience (Vennin et al., 2022).

Second, the mapping of connectivity estimates to regional effects, allows one to account for atrophy at a regional level (Figure S2). This ensures that the more pronounced functional resilience profile in presymptomatic carriers is independent of regional atrophy, rather than only adjusted for global atrophy (Rittman et al., 2019; Tsvetanov et al., 2020). It is worth noting that this framework facilitates the integration of connectomes (e.g. structural and functional connectivity) with regional effects from multiple modalities, including grey matter, perfusion, cerebrovascular reactivity, metabolic rates of glucose, oxygen, and aerobic glycolysis (Vaishnavi et al., 2010) and receptor and neurotransmitter maps (Hansen et al., 2022), in addition transcriptomic patterns as demonstrated above. We propose that by adopting this comprehensive, multimodal perspective, one can advance insights and track the complex multilevel changes preceding clinical symptoms in neurodegenerative diseases such as FTD (Jack et al., 2018; Rosen et al., 2020; Staffaroni et al., 2022). This is especially important where the presymptomatic stage of active disease offers a critical window of opportunity for preventive disease-modifying treatment (Lian et al., 2024).

Third, our approach is based on data-driven decomposition of functional connectivity maps connectivity to higher-dimensional space of neuronal effects (Bethlehem et al., 2020; Margulies et al., 2016). This mitigates the bias from vascular contributions to fMRI BOLD signals (Drew, 2019; Logothetis, 2008), which can be differentially affected by ageing (Henson et al., 2024; Tsvetanov et al., 2021a, 2015), and systemic-to-vascular interactions (Kancheva et al., 2024; Millar et al., 2020; Tsvetanov et al., 2022). By projecting the fMRI BOLD signal into a set of functional components, we isolate the neuronal components of the signal, while filtering physiological noise to other components (Tsvetanov et al., 2021b). This is valuable even in the absence of explicit measures of vascular or neuronal signals (Tsvetanov et al., 2021b).

Despite the large size of the GENFI cohort, this study has several limitations. Genetic groups were not analysed separately due to potential subgroup imbalances and lower statistical power. Similarly, environmental factors and genetic modifiers were not fully addressed (Premi et al., 2017; Stern, 2012). While efforts were made to harmonise data acquisition and reduce site-specific effects, residual scanner variance cannot be ruled out. The cross-sectional design limits causal inferences about progression of atrophy and resilience effects. Moreover, the study did not differentiate between grey matter atrophy and structural connectivity effects on functional integration, which may be essential as cognition’s dependence on functional integration is partly conditional on structural integrity, at least in ageing (Liu et al., 2023). Finally, the findings are limited to autosomal dominant FTD and may not generalise to sporadic FTD or may generalise to other forms of dementia. These limitations highlight the need for global initiatives to create larger, longitudinal FTD cohorts, with more diverse neuroimaging measures (Russell, 2022) to better understand gene-specific effects, environmental and genetic moderators, and the causal role of network integration in cognitive preservation and years of onset across different FTD subtypes and in relation to other dementias.

In conclusion, the multivariate data-driven approach reveals that brain functional integration provides resilience, allowing presymptomatic carriers to maintain cognitive performance despite brain atrophy. Localised to cognitive control networks, this resilience is associated with transcriptomic profiles for glial support. This multivariate approach to whole-brain brain function in relation to cognition, genetics and modifiers is well-suited to study the impact of multiple risk factors on neurodegeneration biomarkers before clinical symptoms appear. Our findings have implications for presymptomatic therapy trials, which are likely to rely initially on surrogate markers of brain health rather than clinical end points. Furthermore, our findings open new avenues for therapeutic strategies aimed at enhancing resilience, with targeted interventions for preserving cognitive function in the face of neurodegeneration.

## Data Availability

https://portal.dementiasplatform.uk/Apply

## 5. Acknowledgements

K.A.T. was supported by Fellowship awards from the Guarantors of Brain (G101149) and the Alzheimer’s Society, UK (Grant number 602).

J.B.R was supported by the NIHR Cambridge Biomedical Research Centre (NIHR203312: BRC-1215-20014), NIHR funding to the NIHR BioResource (RG94028 & RG85445), Wellcome Trust (220258), Medical Research Council (SUAG/051G101400; SUAG/010 RG91365; MC_UU_00030/14 and MR/T033371/1), the Holt Fellowship and by the Addenbrookes Charitable Trust. We thank NIHR BioResource volunteers for their participation, and gratefully acknowledge NIHR BioResource centres, NHS Trusts and staff for their contribution. The views expressed are those of the author(s) and not necessarily those of the NHS or the NIHR.

J.C.V.S., L.C.J. and H.S. are supported by the Dioraphte Foundation grant 09-02-03-00, Association for Frontotemporal Dementias Research Grant 2009, Netherlands Organization for Scientific Research grant HCMI 056-13-018, ZonMw Memorabel (Deltaplan Dementie, project number 733 051 042), ZonMw Onderzoeksprogramma Dementie (YOD-INCLUDED, project number10510032120002), EU Joint Programme-Neurodegenerative Disease Research-GENFI-PROX, Alzheimer Nederland and the Bluefield Project.

C.G. received funding from EU Joint Programme-Neurodegenerative Disease Research-Prefrontals Vetenskapsrådet Dnr 529-2014-7504, EU Joint Programme-Neurodegenerative Disease Research-GENFI-PROX, Vetenskapsrådet 2019-0224, Vetenskapsrådet 2015-02926, Vetenskapsrådet 2018-02754, the Swedish FTD Inititative-Schörling Foundation, Alzheimer Foundation, Brain Foundation, Dementia Foundation and Region Stockholm ALF-project.

C.G. is supported by the Swedish Frontotemporal Dementia Initiative Schörling Foundation; Vetenskapsrådet (Swedish Research Council) JPND Prefrontals, 2015–02926, 2018–02754, and JPND-GENFI-PROX 2019-02248; Swedish Alzheimer Foundation, ALF-project Region Stockholm, Karolinska Institutet Doctoral Funding, KI Strat-Neuro, Swedish Dementia Foundation, and Swedish BrainFoundation.

D.G. received support from the EU Joint Programme—Neurodegenerative Disease Research and the Italian Ministry of Health (PreFrontALS) grant 733051042.

R.V. has received funding from the Mady Browaeys Fund for Research into Frontotemporal Dementia (Mady Browaeys Fonds voor Onderzoek naar Frontotemporale Degeneratie).

J.L. received funding for this work by the Deutsche Forschungsgemeinschaft German Research Foundation under Germany’s Excellence Strategy within the framework of the Munich Cluster for Systems Neurology (EXC 2145 SyNergy—ID 390857198).

E.F. has received funding from a Canadian Institute of Health Research grant #327387. MM was, in part, funded by the UK Medical Research Council, the Italian Ministry of

Health and the Canadian Institutes of Health Research as part of a Centres of Excellence in Neurodegeneration grant, and also Canadian Institutes of Health Research operating grants (Grant #s: MOP-371851 and PJT-175242) and funding from the Weston Brain Institute to Mario Masellis.

FM is supported by the Tau Consortium and has received funding from the Carlos III Health Institute (PI19/01637).

Several authors of this publication (J.C.V.S., M.S., R.V., A.d.M., M.O., R.V., J.D.R.) are members of the European Reference Network for Rare Neurological Diseases (ERN-RND) - Project ID No 739510.

This work was also supported by the EU Joint Programme—Neurodegenerative Disease Research GENFI-PROX grant [2019-02248; to J.D.R., M.O., B.B., C.G., J.C.V.S. and M.S.

RS-V was funded at the Hospital Clinic de Barcelona by Instituto de Salud Carlos III, Spain (grant code PI20/00448 to RSV) and Fundació Marató TV3, Spain (grant code 20143810 to RSV).

RL is supported by the Canadian Institutes of Health Research and theChaire de Recherche sur les Aphasies Primaires Progressives Fondation Famille Lemaire.

This work was funded by Mady Browaeys Fonds voor Onderzoek naar Frontotemporale Degeneratie.

SD receives salary funding from the Fonds de Recherche du Québec - Santé. This research was undertaken thanks in part to funding from the Canada First Research Excellence Fund, awarded to McGill University for the Healthy Brains, Healthy Lives initiative.

This work was supported by ANR-PRTS PREV-DemAls, PHRC PREDICT-PGRN and several authors of this publication are members of the European Reference Network for Rare Neurological Diseases - Project ID No 739510.

This work was supported by the JPND grant “GENFI-prox” (by DLR/BMBF to M.S, joint with J.R., JvS, M.O., B.B. and C.G.).

FM is supported by the Tau Consortium and has received funding from the Carlos III Health Institute (PI19/01637).

BB is supported by JPND grant “GENFI-prox” (2019-02248).

JDR is supported by the Miriam Marks Brain Research UK Senior Fellowship and has received funding from an MRC Clinician Scientist Fellowship (MR/M008525/1) and the NIHR Rare Disease Translational Research Collaboration (BRC149/NS/MH). This work was also supported by the MRC UK GENFI grant (MR/M023664/1), the Bluefield Project and the JPND GENFI-PROX grant (2019-02248).

For the purpose of open access, the author has applied a CC BY public copyright licence to any Author Accepted Manuscript version arising from this submission.

## 6. Funding

NIHR Cambridge Biomedical Research Centre (NIHR203312: BRC-1215-20014)

NIHR funding to the NIHR BioResource (RG94028)

NIHR funding to the NIHR BioResource (RG85445)

Wellcome Trust (220258)

Medical Research Council (SUAG/010 RG91365)

Medical Research Council (SUAG/051G101400)

Holt Fellowship

Addenbrookes Charitable Trust

Guarantors of Brain (G101149)

Alzheimer’s Society (grant number 602)

Race Against Dementia Alzheimer’s Research UK (ARUK-RADF2021A-010)

## 7. Competing interests

All authors have no conflicts of interest. Untreated to this there are several disclosures.

JBR is a non-remunerated trustee of the Guarantors of Brain, Darwin College, and the PSP Association; he provides consultancy to Alzheimer Research UK, Asceneuron, Alector, Biogen, CuraSen, CumulusNeuro, UCB, SV Health, and Wave, and has research grants from AZ-Medimmune, Janssen, Lilly as industry partners in the Dementias Platform UK.

Prof. Lebouvier receives consultancy fees from Roche, Lilly, Biogen, and Eisai, which are directed entirely to his institution.

M.M. has acted as a consultant for Astex Pharmaceuticals.

## 8. Appendix

The GENFI Consortium Author List:

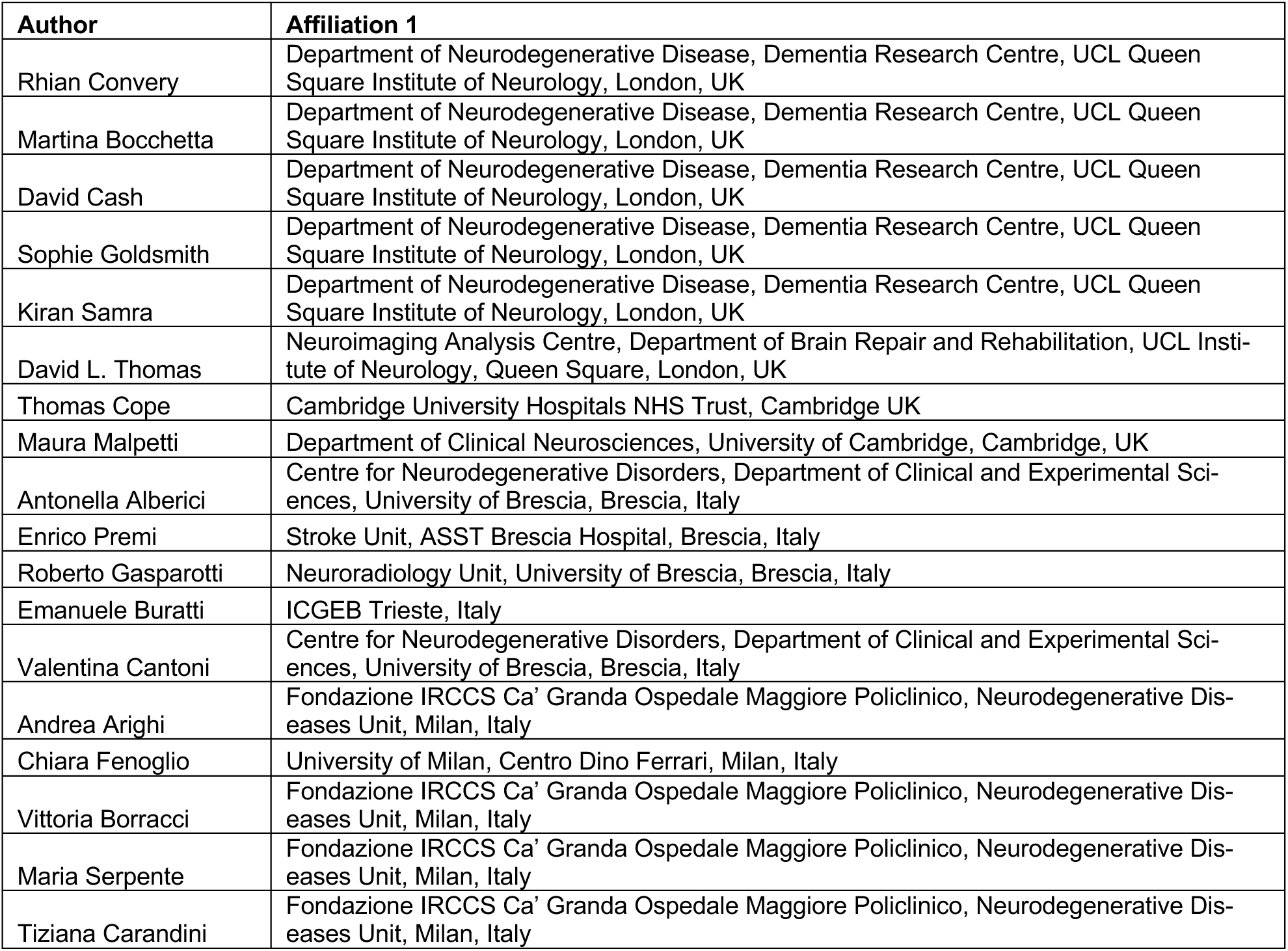
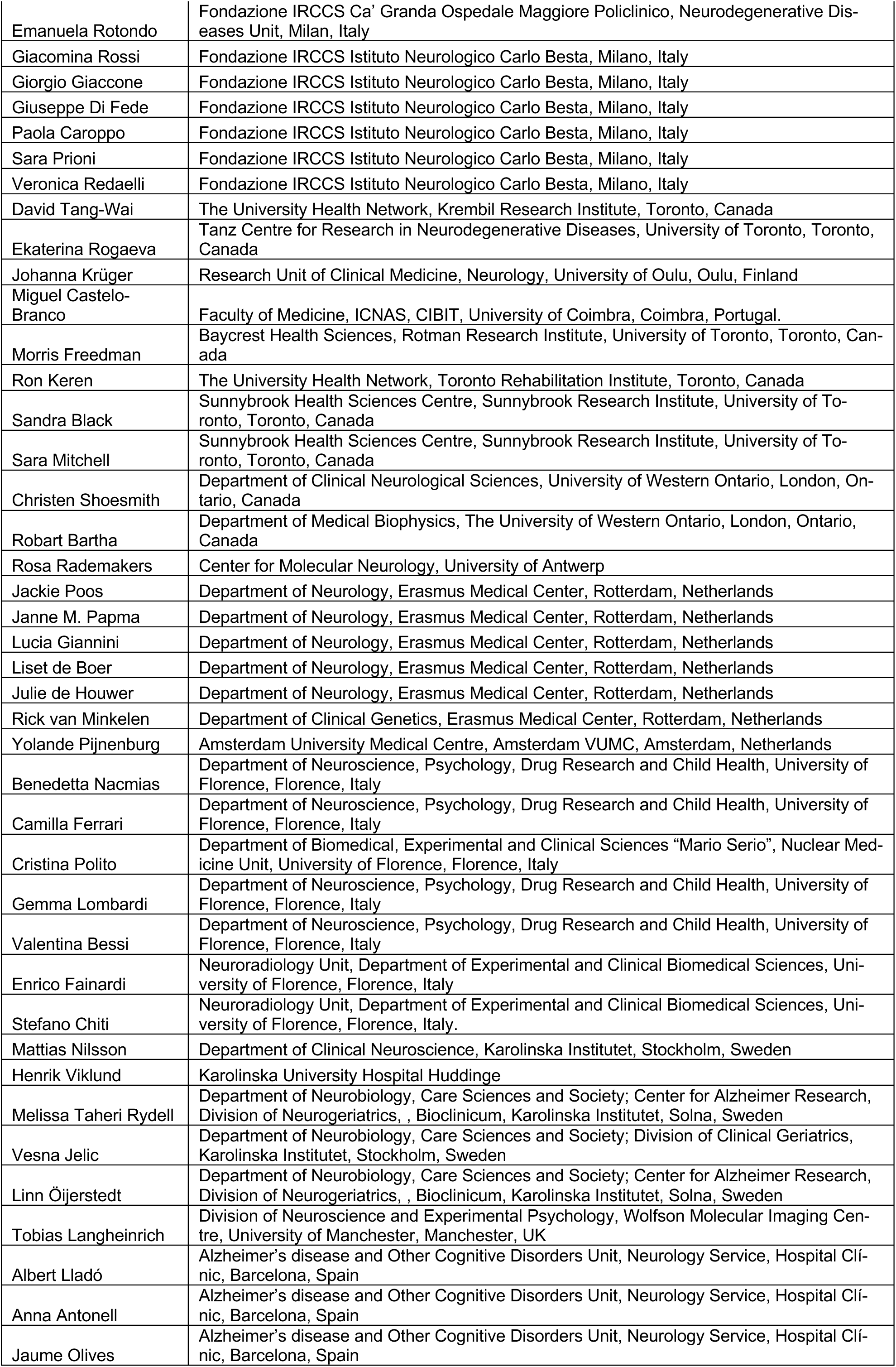
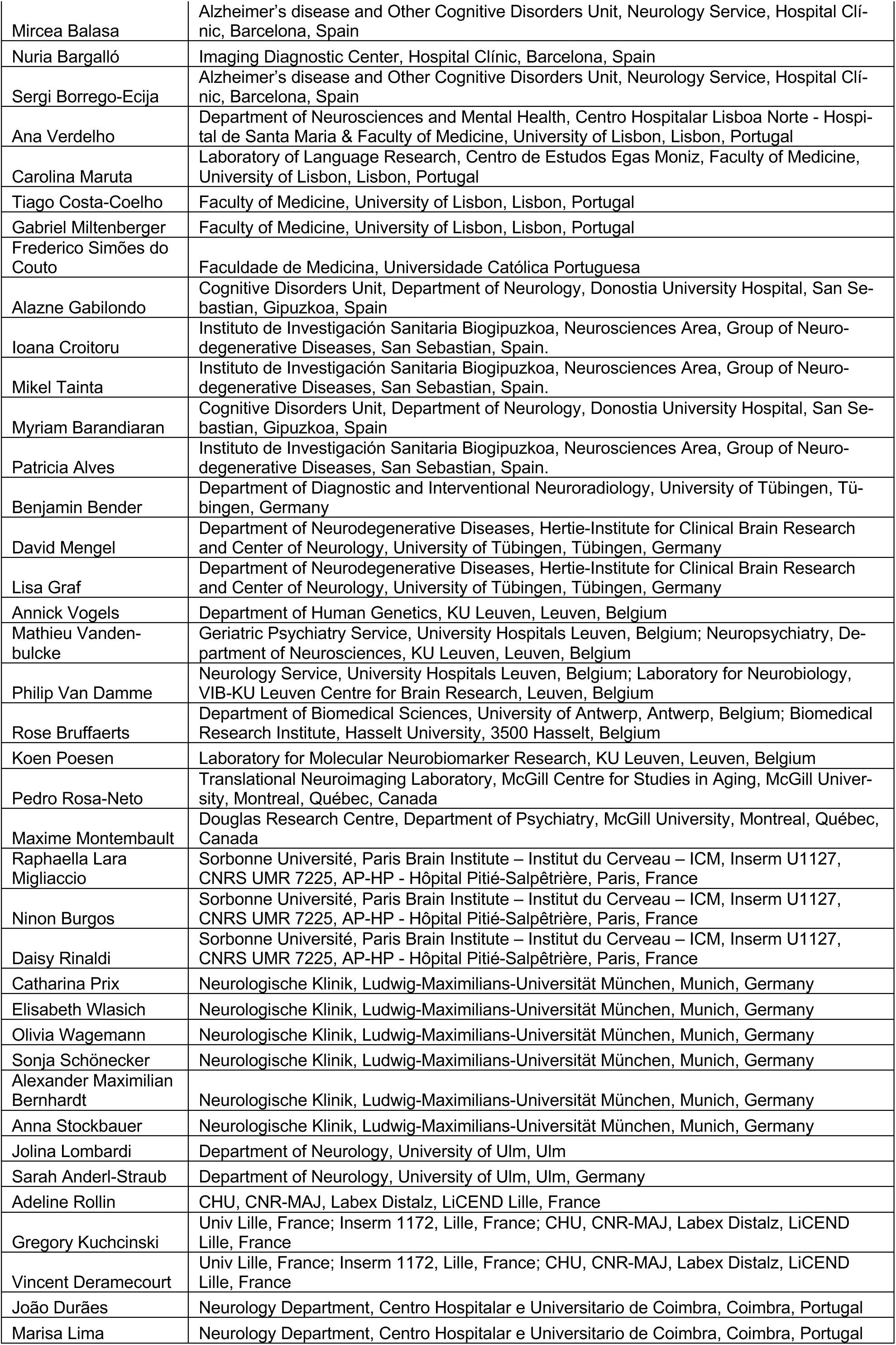
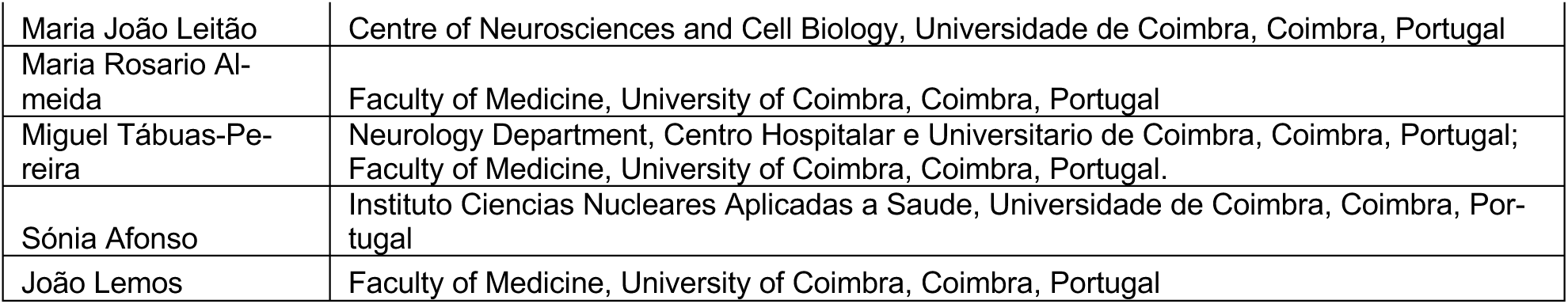

| Author | Affiliation 1 |
| --- | --- |
| Rhian Convery | Department of Neurodegenerative Disease, Dementia Research Centre, UCL Queen Square Institute of Neurology, London, UK |
| Martina Bocchetta | Department of Neurodegenerative Disease, Dementia Research Centre, UCL Queen Square Institute of Neurology, London, UK |
| David Cash | Department of Neurodegenerative Disease, Dementia Research Centre, UCL Queen Square Institute of Neurology, London, UK |
| Sophie Goldsmith | Department of Neurodegenerative Disease, Dementia Research Centre, UCL Queen Square Institute of Neurology, London, UK |
| Kiran Samra | Department of Neurodegenerative Disease, Dementia Research Centre, UCL Queen Square Institute of Neurology, London, UK |
| David L. Thomas | Neuroimaging Analysis Centre, Department of Brain Repair and Rehabilitation, UCL Institute of Neurology, Queen Square, London, UK |
| Thomas Cope | Cambridge University Hospitals NHS Trust, Cambridge UK |
| Maura Malpetti | Department of Clinical Neurosciences, University of Cambridge, Cambridge, UK |
| Antonella Alberici | Centre for Neurodegenerative Disorders, Department of Clinical and Experimental Sciences, University of Brescia, Brescia, Italy |
| Enrico Premi | Stroke Unit, ASST Brescia Hospital, Brescia, Italy |
| Roberto Gasparotti | Neuroradiology Unit, University of Brescia, Brescia, Italy |
| Emanuele Buratti | ICGEB Trieste, Italy |
| Valentina Cantoni | Centre for Neurodegenerative Disorders, Department of Clinical and Experimental Sciences, University of Brescia, Brescia, Italy |
| Andrea Arighi | Fondazione IRCCS Ca' Granda Ospedale Maggiore Policlinico, Neurodegenerative Diseases Unit, Milan, Italy |
| Chiara Fenoglio | University of Milan, Centro Dino Ferrari, Milan, Italy |
| Vittoria Borracci | Fondazione IRCCS Ca' Granda Ospedale Maggiore Policlinico, Neurodegenerative Diseases Unit, Milan, Italy |
| Maria Serpente | Fondazione IRCCS Ca' Granda Ospedale Maggiore Policlinico, Neurodegenerative Diseases Unit, Milan, Italy |
| Tiziana Carandini | Fondazione IRCCS Ca' Granda Ospedale Maggiore Policlinico, Neurodegenerative Diseases Unit, Milan, Italy |
| Emanuela Rotondo | Fondazione IRCCS Ca' Granda Ospedale Maggiore Policlinico, Neurodegenerative Diseases Unit, Milan, Italy |
| Giacomina Rossi | Fondazione IRCCS Istituto Neurologico Carlo Besta, Milano, Italy |
| Giorgio Giaccone | Fondazione IRCCS Istituto Neurologico Carlo Besta, Milano, Italy |
| Giuseppe Di Fede | Fondazione IRCCS Istituto Neurologico Carlo Besta, Milano, Italy |
| Paola Caroppo | Fondazione IRCCS Istituto Neurologico Carlo Besta, Milano, Italy |
| Sara Prioni | Fondazione IRCCS Istituto Neurologico Carlo Besta, Milano, Italy |
| Veronica Redaelli | Fondazione IRCCS Istituto Neurologico Carlo Besta, Milano, Italy |
| David Tang-Wai | The University Health Network, Krembil Research Institute, Toronto, Canada |
| Ekaterina Rogaeva | Tanz Centre for Research in Neurodegenerative Diseases, University of Toronto, Toronto, Canada |
| Johanna Krüger | Research Unit of Clinical Medicine, Neurology, University of Oulu, Oulu, Finland |
| Miguel Castelo-Branco | Faculty of Medicine, ICNAS, CIBIT, University of Coimbra, Coimbra, Portugal. |
| Morris Freedman | Baycrest Health Sciences, Rotman Research Institute, University of Toronto, Toronto, Canada |
| Ron Keren | The University Health Network, Toronto Rehabilitation Institute, Toronto, Canada |
| Sandra Black | Sunnybrook Health Sciences Centre, Sunnybrook Research Institute, University of Toronto, Toronto, Canada |
| Sara Mitchell | Sunnybrook Health Sciences Centre, Sunnybrook Research Institute, University of Toronto, Toronto, Canada |
| Christen Shoesmith | Department of Clinical Neurological Sciences, University of Western Ontario, London, Ontario, Canada |
| Robart Bartha | Department of Medical Biophysics, The University of Western Ontario, London, Ontario, Canada |
| Rosa Rademakers | Center for Molecular Neurology, University of Antwerp |
| Jackie Poos | Department of Neurology, Erasmus Medical Center, Rotterdam, Netherlands |
| Janne M. Papma | Department of Neurology, Erasmus Medical Center, Rotterdam, Netherlands |
| Lucia Giannini | Department of Neurology, Erasmus Medical Center, Rotterdam, Netherlands |
| Liset de Boer | Department of Neurology, Erasmus Medical Center, Rotterdam, Netherlands |
| Julie de Houwer | Department of Neurology, Erasmus Medical Center, Rotterdam, Netherlands |
| Rick van Minkelen | Department of Clinical Genetics, Erasmus Medical Center, Rotterdam, Netherlands |
| Yolande Pijnenburg | Amsterdam University Medical Centre, Amsterdam VUMC, Amsterdam, Netherlands |
| Benedetta Nacmias | Department of Neuroscience, Psychology, Drug Research and Child Health, University of Florence, Florence, Italy |
| Camilla Ferrari | Department of Neuroscience, Psychology, Drug Research and Child Health, University of Florence, Florence, Italy |
| Cristina Polito | Department of Biomedical, Experimental and Clinical Sciences "Mario Serio", Nuclear Medicine Unit, University of Florence, Florence, Italy |
| Gemma Lombardi | Department of Neuroscience, Psychology, Drug Research and Child Health, University of Florence, Florence, Italy |
| Valentina Bessi | Department of Neuroscience, Psychology, Drug Research and Child Health, University of Florence, Florence, Italy |
| Enrico Fainardi | Neuroradiology Unit, Department of Experimental and Clinical Biomedical Sciences, University of Florence, Florence, Italy |
| Stefano Chiti | Neuroradiology Unit, Department of Experimental and Clinical Biomedical Sciences, University of Florence, Florence, Italy. |
| Mattias Nilsson | Department of Clinical Neuroscience, Karolinska Institutet, Stockholm, Sweden |
| Henrik Viklund | Karolinska University Hospital Huddinge |
| Melissa Taheri Rydell | Department of Neurobiology, Care Sciences and Society; Center for Alzheimer Research, Division of Neurogeriatrics, , Bioclinicum, Karolinska Institutet, Solna, Sweden |
| Vesna Jelic | Department of Neurobiology, Care Sciences and Society; Division of Clinical Geriatrics, Karolinska Institutet, Stockholm, Sweden |
| Linn Öijerstedt | Department of Neurobiology, Care Sciences and Society; Center for Alzheimer Research, Division of Neurogeriatrics, , Bioclinicum, Karolinska Institutet, Solna, Sweden |
| Tobias Langheinrich | Division of Neuroscience and Experimental Psychology, Wolfson Molecular Imaging Centre, University of Manchester, Manchester, UK |
| Albert Lladó | Alzheimer's disease and Other Cognitive Disorders Unit, Neurology Service, Hospital Clínic, Barcelona, Spain |
| Anna Antonell | Alzheimer's disease and Other Cognitive Disorders Unit, Neurology Service, Hospital Clínic, Barcelona, Spain |
| Jaume Olives | Alzheimer's disease and Other Cognitive Disorders Unit, Neurology Service, Hospital Clínic, Barcelona, Spain |
| Mircea Balasa | Alzheimer's disease and Other Cognitive Disorders Unit, Neurology Service, Hospital Clínic, Barcelona, Spain |
| Nuria Bargalló | Imaging Diagnostic Center, Hospital Clínic, Barcelona, Spain |
| Sergi Borrego-Ecija | Alzheimer's disease and Other Cognitive Disorders Unit, Neurology Service, Hospital Clínic, Barcelona, Spain |
| Ana Verdelho | Department of Neurosciences and Mental Health, Centro Hospitalar Lisboa Norte - Hospital de Santa Maria & Faculty of Medicine, University of Lisbon, Lisbon, Portugal |
| Carolina Maruta | Laboratory of Language Research, Centro de Estudos Egas Moniz, Faculty of Medicine, University of Lisbon, Lisbon, Portugal |
| Tiago Costa-Coelho | Faculty of Medicine, University of Lisbon, Lisbon, Portugal |
| Gabriel Miltenberger | Faculty of Medicine, University of Lisbon, Lisbon, Portugal |
| Frederico Simões do Couto | Faculdade de Medicina, Universidade Católica Portuguesa |
| Alazne Gabilondo | Cognitive Disorders Unit, Department of Neurology, Donostia University Hospital, San Sebastian, Gipuzkoa, Spain |
| Ioana Croitoru | Instituto de Investigación Sanitaria Biogipuzkoa, Neurosciences Area, Group of Neurodegenerative Diseases, San Sebastian, Spain. |
| Mikel Tainta | Instituto de Investigación Sanitaria Biogipuzkoa, Neurosciences Area, Group of Neurodegenerative Diseases, San Sebastian, Spain. |
| Myriam Barandiaran | Cognitive Disorders Unit, Department of Neurology, Donostia University Hospital, San Sebastian, Gipuzkoa, Spain |
| Patricia Alves | Instituto de Investigación Sanitaria Biogipuzkoa, Neurosciences Area, Group of Neurodegenerative Diseases, San Sebastian, Spain. |
| Benjamin Bender | Department of Diagnostic and Interventional Neuroradiology, University of Tübingen, Tübingen, Germany |
| David Mengel | Department of Neurodegenerative Diseases, Hertie-Institute for Clinical Brain Research and Center of Neurology, University of Tübingen, Tübingen, Germany |
| Lisa Graf | Department of Neurodegenerative Diseases, Hertie-Institute for Clinical Brain Research and Center of Neurology, University of Tübingen, Tübingen, Germany |
| Annick Vogels | Department of Human Genetics, KU Leuven, Leuven, Belgium |
| Mathieu Vandenbulcke | Geriatric Psychiatry Service, University Hospitals Leuven, Belgium; Neuropsychiatry, Department of Neurosciences, KU Leuven, Leuven, Belgium |
| Philip Van Damme | Neurology Service, University Hospitals Leuven, Belgium; Laboratory for Neurobiology, VIB-KU Leuven Centre for Brain Research, Leuven, Belgium |
| Rose Bruffaerts | Department of Biomedical Sciences, University of Antwerp, Antwerp, Belgium; Biomedical Research Institute, Hasselt University, 3500 Hasselt, Belgium |
| Koen Poesen | Laboratory for Molecular Neurobiomarker Research, KU Leuven, Leuven, Belgium |
| Pedro Rosa-Neto | Translational Neuroimaging Laboratory, McGill Centre for Studies in Aging, McGill University, Montreal, Québec, Canada |
| Maxime Montembault | Douglas Research Centre, Department of Psychiatry, McGill University, Montreal, Québec, Canada |
| Raphaella Lara Migliaccio | Sorbonne Université, Paris Brain Institute – Institut du Cerveau – ICM, Inserm U1127, CNRS UMR 7225, AP-HP - Hôpital Pitié-Salpêtrière, Paris, France |
| Ninon Burgos | Sorbonne Université, Paris Brain Institute – Institut du Cerveau – ICM, Inserm U1127, CNRS UMR 7225, AP-HP - Hôpital Pitié-Salpêtrière, Paris, France |
| Daisy Rinaldi | Sorbonne Université, Paris Brain Institute – Institut du Cerveau – ICM, Inserm U1127, CNRS UMR 7225, AP-HP - Hôpital Pitié-Salpêtrière, Paris, France |
| Catharina Prix | Neurologische Klinik, Ludwig-Maximilians-Universität München, Munich, Germany |
| Elisabeth Wlasich | Neurologische Klinik, Ludwig-Maximilians-Universität München, Munich, Germany |
| Olivia Wagemann | Neurologische Klinik, Ludwig-Maximilians-Universität München, Munich, Germany |
| Sonja Schönecker | Neurologische Klinik, Ludwig-Maximilians-Universität München, Munich, Germany |
| Alexander Maximilian Bernhardt | Neurologische Klinik, Ludwig-Maximilians-Universität München, Munich, Germany |
| Anna Stockbauer | Neurologische Klinik, Ludwig-Maximilians-Universität München, Munich, Germany |
| Jolina Lombardi | Department of Neurology, University of Ulm, Ulm |
| Sarah Anderl-Straub | Department of Neurology, University of Ulm, Ulm, Germany |
| Adeline Rollin | CHU, CNR-MAJ, Labex Distalz, LiCEND Lille, France |
| Gregory Kuchcinski | Univ Lille, France; Inserm 1172, Lille, France; CHU, CNR-MAJ, Labex Distalz, LiCEND Lille, France |
| Vincent Deramecourt | Univ Lille, France; Inserm 1172, Lille, France; CHU, CNR-MAJ, Labex Distalz, LiCEND Lille, France |
| João Durães | Neurology Department, Centro Hospitalar e Universitario de Coimbra, Coimbra, Portugal |
| Marisa Lima | Neurology Department, Centro Hospitalar e Universitario de Coimbra, Coimbra, Portugal |
| Maria João Leitão | Centre of Neurosciences and Cell Biology, Universidade de Coimbra, Coimbra, Portugal |
| Maria Rosario Almeida | Faculty of Medicine, University of Coimbra, Coimbra, Portugal |
| Miguel Tábuas-Pereira | Neurology Department, Centro Hospitalar e Universitário de Coimbra, Coimbra, Portugal; Faculty of Medicine, University of Coimbra, Coimbra, Portugal. |
| Sónia Afonso | Instituto Ciencias Nucleares Aplicadas a Saude, Universidade de Coimbra, Coimbra, Portugal |
| João Lemos | Faculty of Medicine, University of Coimbra, Coimbra, Portugal |

## Notes

### Author Declarations

The study was given a favorable opinion by the Cambridge 2 Research Ethics Committee REC 17/EE/0032 IRAS ID 204052.

