## Supplemental Information for "Cellular signatures of functional resilience in presymptomatic frontotemporal dementia"

Supplementary Information: Neurotransmitter and cellular signatures of functional resilience in presymptomatic frontotemporal dementia

### 1. Supplementary tables

### 2. Supplementary Figures and Analyses

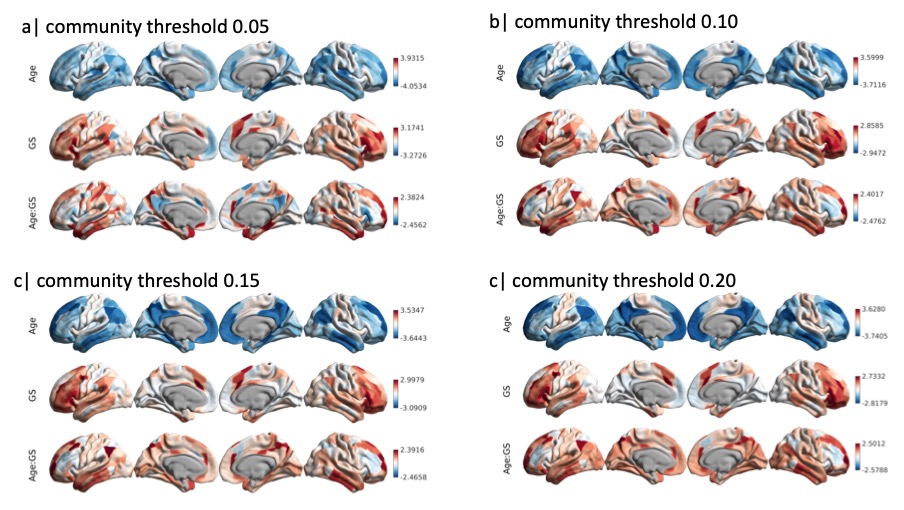

Figure S 1. **Functional integration effects across four threshold levels for selecting communities.** Age, gene status (GS, presymptomatic versus non-carriers) and their interaction (Age:GS) effects on functional integration across four different threshold levels used to define functional communities across the functional gradients.

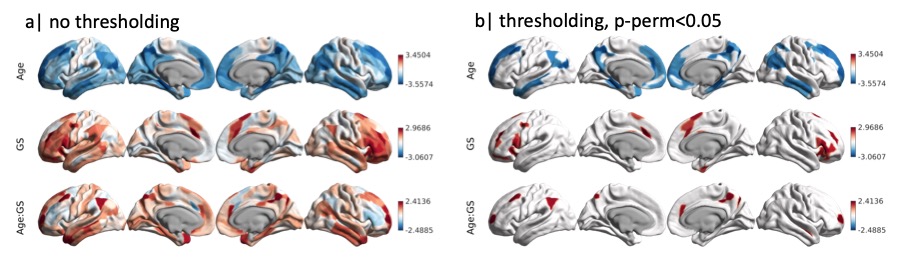

Figure S 2. **Functional integration effects, accounting for cortical thickness.** Age, gene status (GS, presymptomatic versus non-carriers) and their interaction (Age:GS) effects on functional integration in a model accounting for cortical thickness in addition to other covariates of no interest (sex, scanner site, head motion); a| without thresholding for for more complete description of the spatial representation and b| with thresholding at p-perm<0.05, based on 10000 permutations.

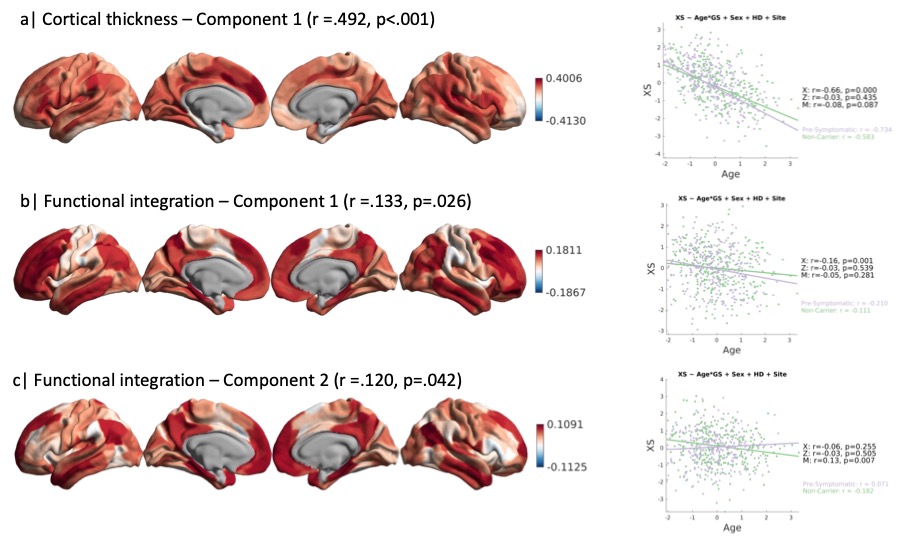

Figure S 3.**Canonical correlation analysis between group effects and brain metrics** (cortical thickness, a, and functional integration, b-c): (a) The first, and only, significant component indicating loss in cortical thickness was exclusively expressed by age. Scatter plot visualises individuals’ expression of this profile as a function of age for both gene status groups, separately (note, lower values indicate weaker expression of the profile, i.e. loss of cortical thickness). (b) The first, of two, significant components indicating loss in functional integrity was exclusively expressed by age. (c) The second significant component indicated a profile of loss of functional integrity that was expressed exclusively by the interaction term Age:Gene Status. Scatter plot indicates that with age non-carriers express less strongly the profile, while presymptomatic carriers maintain functional integrity with increasing age, consistent with the effects observed using a univariate approach.

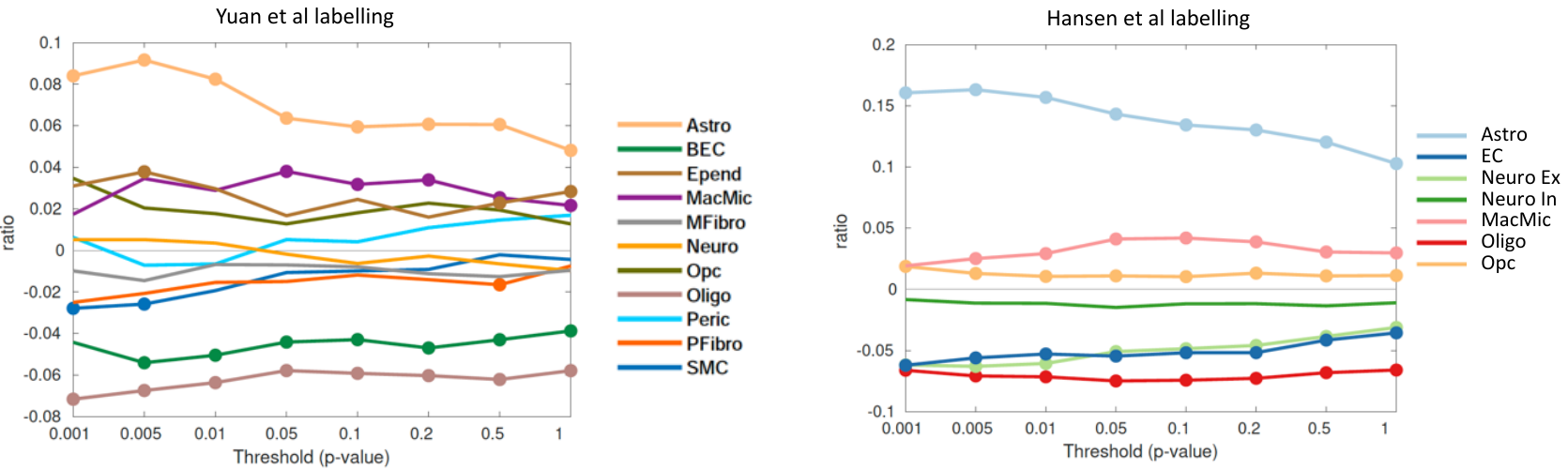

Figure S 4. Spatial correspondence between functional resilience map expressed by functional resilience map, i.e. Canonical Variate 2, and cell-type decomposition based on two labelling schemes: Yuan et al (left panel, (Yang et al., 2022)) and Hansen et al (right panel, (Hansen et al., 2021)). Specific cell-type expression in functional resilience map constructed from the most significant loadings, ranging from p-spin<0.001 to 1. Each point represents the difference between the ratio of genes in each gene set preferentially expressed in a cell-type and the mean null ratio, computed from null distribution of random gene sets (10.000 permutations). Curves above zero indicate overexpression and curves below zero indicate underexpression. Circles demonstrate significance. Astro – astrocytes, BEC – brain endothelial cells, Epend – ependymal, MacMic – macrophage/microglia, MFibro – meningeal fibroblast, Neuro – neuron, Opc – oligodendrocyte precursors, Oligo – oligodendrocytes, Peri – pericytes, PFibro – perivascular fibrblast, SMC – smooth muscle cells, EC – endothelial cells, Neuro Ex – excitatory neurons, Neuro In – inhibitory neurons.
